# Statin use in relation to intraocular pressure, glaucoma, and ocular coherence tomography parameters in the UK Biobank

**DOI:** 10.1101/2021.12.12.21267685

**Authors:** Jihye Kim, Marianne T. Kennedy Neary, Hugues Aschard, Mathew M. Palakkamanil, Ron Do, Janey L. Wiggs, Anthony P. Khawaja, Louis R. Pasquale, Jae H. Kang, for the Modifiable Risk Factors for Glaucoma Collaboration

**Affiliations:** Department of Epidemiology, Harvard T.H. Chan School of Public Health, Boston, Massachusetts, USA; University of Manchester & Manchester Royal Eye Hospital, Manchester, UK; Institut Pasteur, Université de Paris, Department of Computational Biology, F-75015 Paris, France; Department of Ophthalmology and Visual Science, Dalhousie University, Halifax, Nova Scotia, Canada; Charles Bronfman Institute for Personalized Medicine, Department of Genetics and Genomics, Icahn School of Medicine at Mount Sinai, New York, New York, USA; Department of Ophthalmology, Harvard Medical School, Massachusetts Eye and Ear Infirmary, Boston, Massachusetts, USA; NIHR Biomedical Research Centre at Moorfields Eye Hospital & UCL Institute of Ophthalmology, London, UK; Department of Ophthalmology, Icahn School of Medicine, Mount Sinai, New York, New York, USA; Channing Division of Network Medicine, Brigham and Women’s Hospital, Harvard Medical School, Boston, Massachusetts, USA

**Keywords:** statin, intraocular pressure, glaucoma

## Abstract

**Objective:** To evaluate the relationship between statin use and various glaucoma-related traits.

**Design:** Cross-sectional analysis of UK Biobank data.

**Participants:** We included 118,153 participants (mean age (SD)=56.8 (8.0) years) with data on statin use (5 statin types – 2006-2010) and corneal-compensated IOP measured in 2009-2013). Also, we included 192,283 participants (with 8,982 self-reported glaucoma cases as of 2006-2010) for the glaucoma analyses. After excluding participants with neurodegenerative diseases, 41,638 participants with global macular retinal nerve fiber layer thickness (mRNFL) and 41,547 participants with ganglion cell inner plexiform layer thickness (mGCIPL) measurements in 2009–2010 were available for analysis.

**Method:** We examined associations with statin use utilizing multivariable-adjusted linear regression models for IOP, mRNFL, and mGCIPL and logistic regression models for glaucoma. We assessed whether a 2,673-member polygenic risk score (PRS) identified from a glaucoma multi-trait analysis of genome wide association study (MTAG) modified associations. We performed Mendelian randomization (MR) experiments using 5 gene variants as proxies for the cholesterol-altering effect of statins to investigate associations with various glaucoma-related outcomes.

**Main Outcome and Measures:** IOP; glaucoma; mRNFL; mGCIPL.

**Results:** Statin users had higher unadjusted mean IOP ± SD (16.3 ± 3.9 mm Hg; n = 20,593 participants) than non-users (15.9 ± 3.8 mm Hg; n = 97,560 participants), but in a multivariable-adjusted model, IOP did not differ by statin use (difference = 0.05 mm Hg; 95% CI: -0.02, 0.13; p*=*0.17). Similarly, statin use was not associated with prevalent glaucoma (OR = 1.05; 95% CI: 0.98, 1.13). Statin use was weakly associated with thinner mRNFL (difference = -0.15 microns; 95% CI: -0.28, -0.01; p=0.03) but not with mGCIPL thickness (difference = -0.12 microns; 95% CI: -0.29, 0.05; p=0.17). Among statins, simvastatin and atorvastatin, the two most commonly used statins, were not associated with any glaucoma outcome measures. No association was modified by the glaucoma MTAG PRS (*P*_interaction_*≥*0.16). MR experiments showed no evidence for a causal association between the cholesterol-altering effect of statins and various glaucoma outcomes (inverse weighted variance p*≥*0.14).

**Conclusions:** Statin use was not associated with lower IOP, lower glaucoma prevalence, thicker mRNFL or thicker mGCIPL in the UK Biobank.

---

Statins are a class of lipid-lowering drugs that lower 3-hydroxy-3-methylglutaryl-coenzyme A (HMG-CoA) reductase activity to effectively decrease low density lipoprotein (LDL) cholesterol levels, and they are indicated for the primary prevention of cardiovascular disease.^1, 2^ In the United States, an estimated 39.2 million adults aged 40 years and older were statin users in 2012-2013,^1^ while in a United Kingdom primary care database, the prescription prevalence for statins was 128 per 1000 person-years in 2013.^3^ Given the widespread use of statins, there has been intense interest in their impact on non-cardiovascular outcomes including eye diseases such as diabetic retinopathy^4^ and age related macular degeneration.^5^

Studies of the relation between statin use and glaucoma have yielded mixed results. A meta-analysis of observational studies published in 2016 suggested an inverse relation between statin use and glaucoma.^6^ However, a subsequent large observational study of health professionals illustrated that after careful control for chronological age, the relation between statin use and incident primary open-angle glaucoma (POAG) was null,^7^ and a recent database study in Australia reported an adverse relation between long-term statin use and glaucoma.^8^ Ideally, an observational study of the relation between statins and glaucoma should address exposure misclassification, detection bias, confounding by indication, control for all key covariates and consider possible modification by glaucoma-related genes and Mendelian Randomization (MR)^9^ experiments to address the possibility of a causal effect of statin use in relation to glaucoma.

We used the UK Biobank with its rich repository of questionnaire data, intraocular pressure (IOP) measures, ocular coherence tomography (OCT) parameters, covariate data and genetic biomarkers to address the relation between statin use and various glaucoma-related traits. We also created a genetic instrument variable that served as a proxy for HMG-CoA activity, which reflected long term propensity for having lower LDL levels and has been associated with lower risk of myocardial infarction and death from cardiovascular disease.^10, 11^ We performed MR experiments to assess this instrument in relation to glaucoma-related outcomes.

## Methods

### The UK Biobank

The UK Biobank is a population-based study that continues to collect a broad array of health-related data on over 500,000 participants aged 40-69 years at baseline (2006-2010). We used baseline questionnaire data (http://www.ukbiobank.ac.uk), high throughput genotyping data, ophthalmic data, and OCT imaging data. The UK Biobank was approved by the National Information Governance Board for Health and Social Care and the National Health Service North West Multicenter Research Ethics Committee (reference number 06/MRE08/65). This research was conducted using the UK Biobank Resource under application number 36741. This research study adhered to the tenets of the Declaration of Helsinki.

### Assessment of statin and non-statin hypolipidemic drug use

Trained nurses collected data on prescription drugs or supplements regularly used via in-person interviews at baseline. If a participant indicated on a touchscreen display that they were taking cholesterol lowering drugs, the interviewer recorded the type of medicine used. If the participant indicated they were not using any medications, the interviewer confirmed that the statement was correct. Interviewers did not collect information about dosing or duration of medication use. The statin types used by participants included simvastatin, atorvastatin, rosuvastatin, pravastatin, and fluvastatin. We also analyzed data on use of non-statin hypolipidemic drugs, which included fibric acid agents, bile acid sequestrants, niacin-derivatives, cholesterol absorption inhibitors, omega-3 fats and other miscellaneous antihyperlipidemic agents.

### IOP and glaucoma status ascertainment

In 2009-2013, IOP measurements in both eyes were taken using the Ocular Response Analyzer noncontact tonometer (Reichert Corp., Philadelphia, PA) at 6 locations throughout the UK; for the vast majority of participants, statins were assessed on the same day as IOP measurements (2009-2010) while for a small subset, IOP data was first available in 2012-2013.^12^ Subjects with eye surgery or an eye infection in the prior month were excluded from participating. As a primary outcome, we analyzed corneal-compensated IOP (IOPcc), which is least influenced by measurement artefact due to corneal biomechanics.^13^ We excluded outliers in the extreme top and bottom 0.5 percentiles.^14^ Participants with a history of glaucoma laser or surgical treatment were excluded as their untreated IOP level would not be captured. For those participants on ocular hypotensive therapy, we adopted the convention of adjusting the measured IOP upward by 30%, as we have done in prior publications.^15, 16^ IOPcc measurements were available for right and left eyes on 127,798 and 127,428 participants, respectively; we used the mean of all available right and left eye values. Ultimately, 118,153 eligible participants had complete data to assess the relation between statin use and IOPcc.

At baseline (2006-2010), 9,198 out of 215,562 participants reported that they had glaucoma. Participants completed a touch screen questionnaire and were considered to have glaucoma if in response to the question, “Has a doctor told you that you have any of the following problems with your eyes?”, they chose glaucoma from the menu.

Participants were also considered to have glaucoma if they reported a history of glaucoma surgery or laser on the questionnaire or if they carried an ICD9/10 code for glaucoma (ICD 9: 365.*; ICD10: H40.** (excluding H40.01* and H42.*)). Ultimately, 192,283 participants (including 8,982 glaucoma cases) had sufficient data to assess the relationship between statin use and glaucoma.

### OCT data

Under the auspices of the UK Biobank Eye and Vision Consortium, from 2009-2010, 67,321 individuals underwent macular spectral domain OCT (SD-OCT) imaging as part of the baseline exam.^17^ Loss of retinal ganglion cell (RGC) bodies and their axons are hallmarks of glaucomatous degeneration, and because ∼50% of RGC bodies reside in the macula region, the retinal nerve fiber layer (mRNFL) and ganglion cell inner plexiform layer (mGCIPL) thicknesses in the macula are useful glaucoma-related biomarkers.^18, 19^ The Topcon 3D OCT1000 Mark II was used to complete SD-OCT imaging in a dark room without pupil dilation. The 3-dimensional 6×6 mm^2^ macular volume scan mode (512 A scans per B scan; 128 horizontal B scans in a raster pattern) was employed for imaging. Both eyes were imaged starting with the right eye. The OCT images were stored as downloadable FDA and FDS files from a secure portal. The mRNFL and mGCIPL were segmented using Version 1.6.1.1 of the Topcon Advanced Boundary Segmentation algorithm.^17^ All OCT images with a quality score less than 45 were removed from the data set. Furthermore, as outlined by Khawaja et al,^17^ using various indicator scores that identify blinks, eye motion artefacts, and segmentation failures, we removed participants in the bottom 20th percentile of data quality. The final dataset consisted of 41,638 participants for mRNFL and 41,547 participants for mGCIPL after excluding participants with Alzheimer’s disease, Parkinson’s disease and multiple sclerosis who could have non-glaucomatous reductions in RNFL and GCIPL thicknesses. For mRNFL and mGCIPL measures intended for analyses, we used the mean of all available right and left eye values.

### Genotyping data, glaucoma multi-trait analysis of genome wide association study (MTAG) polygenic risk score and MR experiments

Genetic data on 488,000 participants was generated using two closely related genotyping platforms: the Affymetrix UK BiLEVE Axiom Array, which called genotypes at 807,411 markers on ∼50,000 individuals,^20^ and the Affymetrix UK Biobank Axiom Array, which generated genotypes at 825,925 markers for ∼450,00 individuals. The smaller number of available data partially reflects post-study opt outs and other technical factors. Quality controls and imputation (the determination of genotypes at loci by inference and not by direct genotyping) were performed jointly for these platforms, as previously described.^14^ Specifically, imputation was based on genetic architecture ascertained in the 1000 Genomes Project, UK 10K, and the Haplotype Reference Consortium reference panels. After quality control, 92,693,895 genetic markers of 487,442 participants were available.

For the assessment of whether genetic factors modify the relation between statin use and our various outcomes, we calculated a polygenic risk score (PRS) for each participant using 2,673 independent common single nucleotide polymorphisms (SNPs) associated at *P*≤0.001 for glaucoma from a recent multi-trait analysis of GWAS (MTAG) that included the UK Biobank.^21^ We applied the effect estimates from the MTAG study to generate a glaucoma 2,673-gene variant PRS in the UK Biobank that also predicted earlier age at glaucoma diagnosis, glaucoma progression and need for surgical intervention in an independent Australian case control dataset.^21^ We chose the glaucoma MTAG PRS because it best captures the genetic variation for POAG, which is a complex disease with optic nerve vulnerability across the spectrum of IOP. The PRS was derived using a standard weighted sum of individual SNPs, i.e., 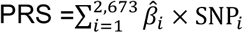 where 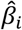 is the estimated effect size of SNP_*j*_ on glaucoma extracted from the aforementioned GWAS.^21^ We normalized the glaucoma PRS with mean of 0 and SD of 1 for analyses.

To address the possibility of a causal effect of statin use in relation to glaucoma, we performed an MR analysis. Analogous to a randomized controlled trial, MR leverages the random allocation of genotypes and allows for the evaluation of associations between genetically predicted levels of a risk factor (e.g., statin use approximated with an instrument incorporating 5 genome-wide significant gene 3-HMG-CoA variants associated with lower serum LDL levels^10^) and genetic predicted propensity for a disease outcome (e.g., glaucoma), which would assist with inferring the causal effect of the risk factor on the outcome.^9^ Briefly, MR leverages genotypic differences that exist at conception. The instrumental variable comprises multiple variants that capture the life-time exposure in a dose-response manner. We conducted the MR study in participants of European descent from the UK Biobank plus other cohorts to optimize the ability to find associations with glaucoma-related traits. Specifically, we used data from the largest available GWAS meta-analyses for POAG (n=216,257)^22^ and for IOP, which includes UK Biobank (n=139,555).^14^ We also chose a GWAS for vertical cup disc ratio (VCDR) ascertained in the International Glaucoma Genetics Consortium (n=23,899) as the assessment of the phenotype in this population was mostly based on scanning laser ophthalmoscopy.^23^ Finally, we picked a subset of the OCT imaging cohort in UK Biobank where both the imaging and genotype quality filters were high for the assessment of both mRNFL and mGCIPL in the same image (n=31,434).^24^ More details regarding the MR experiments can be found in the **Supplemental Methods**.

### Statistical analysis

We compared baseline characteristics of non-statin users and statin users by using mean (SD) difference for continuous variables and differences in frequencies for categorical variables. To examine the main associations between statin use and IOP and between statin use and OCT parameters, we used multiple linear regression models adjusted for covariates. We created 5 nested models to address confounding (especially confounding by indication) and detection bias by including the following covariates: age and age-squared (years; to finely control for age), sex, ethnicity (Caucasian, Black and other), smoking status (never, past and current smoker), number of cigarettes smoked daily among current smokers, alcohol intake frequency (daily or almost daily, 3-4 times a week, 1-2 times a week, 1-3 times a month, special occasions only, never), coffee and tea consumption (cups per day), physical activity (Metabolic Equivalent of Task (MET)-hours/week), Townsend deprivation index (range: -6 to 11; a higher index score indicates more relative poverty for a given residential area), body mass index (BMI) (kg/m^2^), systolic blood pressure (mm Hg), history of diabetes (yes or no) as well as hemoglobin A1c (mmol/mol, to finely control for insulin resistance), history of cardiovascular disease, total serum cholesterol (mmol/l) and triglyceride levels (mmol/l), systemic beta blocker use, use of non-statin hypolipidemic medication and spherical equivalent. To evaluate the relationship between statin use and glaucoma, we conducted multiple logistic regression analyses adjusting for the covariates listed above in the same manner.

To assess whether the glaucoma MTAG PRS modified the relation between statin use and the various outcomes, we tested the significance of adding a multiplicative (MTAG PRS*statin) interaction term in models with the main effects of statin use and the MTAG PRS; in these models, we also included the following multiplicative interaction terms to minimize confounding: total cholesterol*MTAG PRS, cardiovascular disease*MTAG PRS, triglyceride*MTAG PRS, age*MTAG PRS, age^2^*MTAG PRS, and non-statin hypolipidemic drug use*MTAG PRS.^25^

We conducted various sensitivity analyses: (1) analyses excluding those with glaucoma for analyses of IOP and of OCT parameters, (2) a subgroup analysis including only participants with well-controlled serum total cholesterol (< 5.13 mmol/L or 200 mg/dl) and no history of cardiovascular disease, diabetes or beta-blocker use, (3) analysis of participants with complete data on all outcomes (N=41,444), and (4) analyses where the reference group were restricted to people who did not take any hypolipidemic medicines, as opposed to those who were non-statin users.

## Results

The mean age (SD) of the UK Biobank population (n=502,506) at baseline was 56.5 (8.1) years. The population was 82.1% Caucasian and 54.4% were women. The demographics for the specific study population for each analysis described below were similar (**Supplementary Table 1)**.

### Statin use and IOP in the UK Biobank

For the assessment of statin use and IOP, we included 118,153 participants (**Table 1**). Among statin users, the majority (92.7%) reported using either simvastatin or atorvastatin. Statin users were older than non-statin users (61.4 (6.1) versus 55.9 (8.1) years), less likely to be female (38.6% vs 56.7%) and more likely to be using beta-blockers (20.7% vs. 3.0%). Statin users had higher systolic blood pressure, higher BMI and were more likely to report diabetes. The average total cholesterol among statin users was lower (4.7 (0.9) mmol/l) compared to non-statin users (5.9 (1.0) mmol/l; of note, “high cholesterol” is defined as a total level >6.2 mmol/l). The crude average IOP was higher for statin users versus non-statin users (16.3 (3.9) mm Hg vs. 15.9 (3.8) mm Hg). The comparison of statin users, users of non-statin hypolipidemic drugs and non-users of hypolipidemic medicines is provided in **Supplementary Table 2**.

**Table 1.**
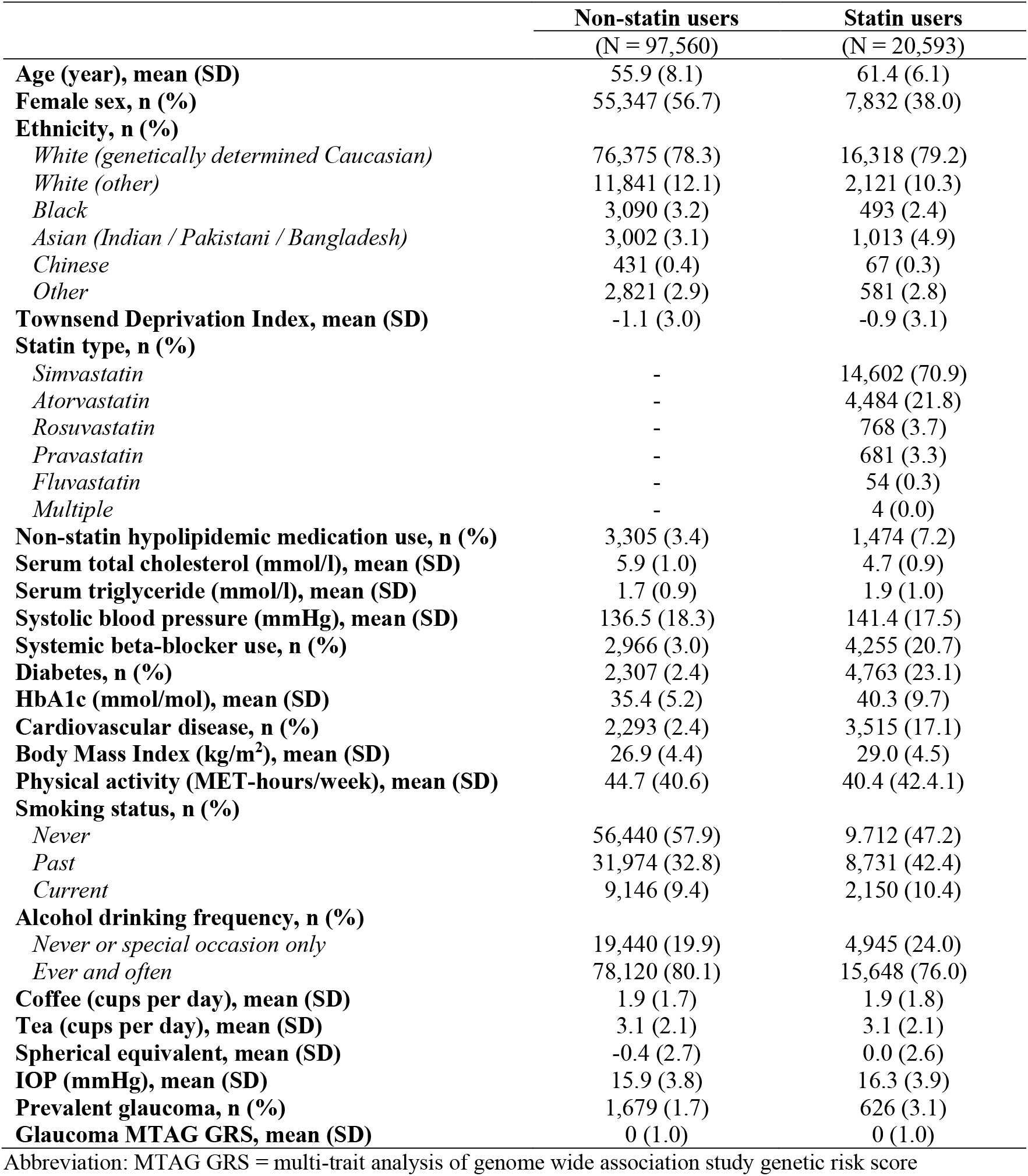
Characteristics of 118,153 UK Biobank participants with intraocular pressure measurements according to statin use at baseline (2006-2010)

|  | **Non-statin users** | **Statin users** |
| --- | --- | --- |
|  | (N = 97,560) | (N = 20,593) |
| **Age (year), mean (SD)** | 55.9 (8.1) | 61.4 (6.1) |
| **Female sex, n (%)** | 55,347 (56.7) | 7,832 (38.0) |
| **Ethnicity, n (%)** |  |  |
| *White (genetically determined Caucasian)* | 76,375 (78.3) | 16,318 (79.2) |
| *White (other)* | 11,841 (12.1) | 2,121 (10.3) |
| *Black* | 3,090 (3.2) | 493 (2.4) |
| *Asian (Indian / Pakistani / Bangladesh)* | 3,002 (3.1) | 1,013 (4.9) |
| *Chinese* | 431 (0.4) | 67 (0.3) |
| *Other* | 2,821 (2.9) | 581 (2.8) |
| **Townsend Deprivation Index, mean (SD)** | -1.1 (3.0) | -0.9 (3.1) |
| **Statin type, n (%)** |  |  |
| *Simvastatin* | - | 14,602 (70.9) |
| *Atorvastatin* | - | 4,484 (21.8) |
| *Rosuvastatin* | - | 768 (3.7) |
| *Pravastatin* | - | 681 (3.3) |
| *Fluvastatin* | - | 54 (0.3) |
| *Multiple* | - | 4 (0.0) |
| **Non-statin hypolipidemic medication use, n (%)** | 3,305 (3.4) | 1,474 (7.2) |
| **Serum total cholesterol (mmol/l), mean (SD)** | 5.9 (1.0) | 4.7 (0.9) |
| **Serum triglyceride (mmol/l), mean (SD)** | 1.7 (0.9) | 1.9 (1.0) |
| **Systolic blood pressure (mmHg), mean (SD)** | 136.5 (18.3) | 141.4 (17.5) |
| **Systemic beta-blocker use, n (%)** | 2,966 (3.0) | 4,255 (20.7) |
| **Diabetes, n (%)** | 2,307 (2.4) | 4,763 (23.1) |
| **HbA1c (mmol/mol), mean (SD)** | 35.4 (5.2) | 40.3 (9.7) |
| **Cardiovascular disease, n (%)** | 2,293 (2.4) | 3,515 (17.1) |
| **Body Mass Index (kg/m<sup>2</sup>), mean (SD)** | 26.9 (4.4) | 29.0 (4.5) |
| **Physical activity (MET-hours/week), mean (SD)** | 44.7 (40.6) | 40.4 (42.4.1) |
| **Smoking status, n (%)** |  |  |
| *Never* | 56,440 (57.9) | 9,712 (47.2) |
| *Past* | 31,974 (32.8) | 8,731 (42.4) |
| *Current* | 9,146 (9.4) | 2,150 (10.4) |
| **Alcohol drinking frequency, n (%)** |  |  |
| *Never or special occasion only* | 19,440 (19.9) | 4,945 (24.0) |
| *Ever and often* | 78,120 (80.1) | 15,648 (76.0) |
| **Coffee (cups per day), mean (SD)** | 1.9 (1.7) | 1.9 (1.8) |
| **Tea (cups per day), mean (SD)** | 3.1 (2.1) | 3.1 (2.1) |
| **Spherical equivalent, mean (SD)** | -0.4 (2.7) | 0.0 (2.6) |
| **IOP (mmHg), mean (SD)** | 15.9 (3.8) | 16.3 (3.9) |
| **Prevalent glaucoma, n (%)** | 1,679 (1.7) | 626 (3.1) |
| **Glaucoma MTAG GRS, mean (SD)** | 0 (1.0) | 0 (1.0) |

Abbreviation: MTAG GRS = multi-trait analysis of genome wide association study genetic risk score

In a basic multivariable model adjusted for age, age-squared, sex, ethnicity, deprivation, spherical equivalent and non-statin hypolipidemic medication use, statin use was associated with lower IOP (Model 1: difference in IOP = -0.13 mm Hg; p=2.9E-05; **Table 2**). This association was slightly attenuated with additional adjustment for cigarette smoking, alcohol use, physical activity, and caffeinated beverage consumption (Model 2: difference = -0.10 mm Hg; p=0.0027). The association remained significant after adjustment for covariates related to metabolic syndrome; namely, BMI, systolic blood pressure, diabetes and HgA1c as well as cardiovascular disease, which is often co-existent with serum lipid disorders (Model 3: difference = -0.12 mm Hg; p=7.4E-04).

**Table 2.**
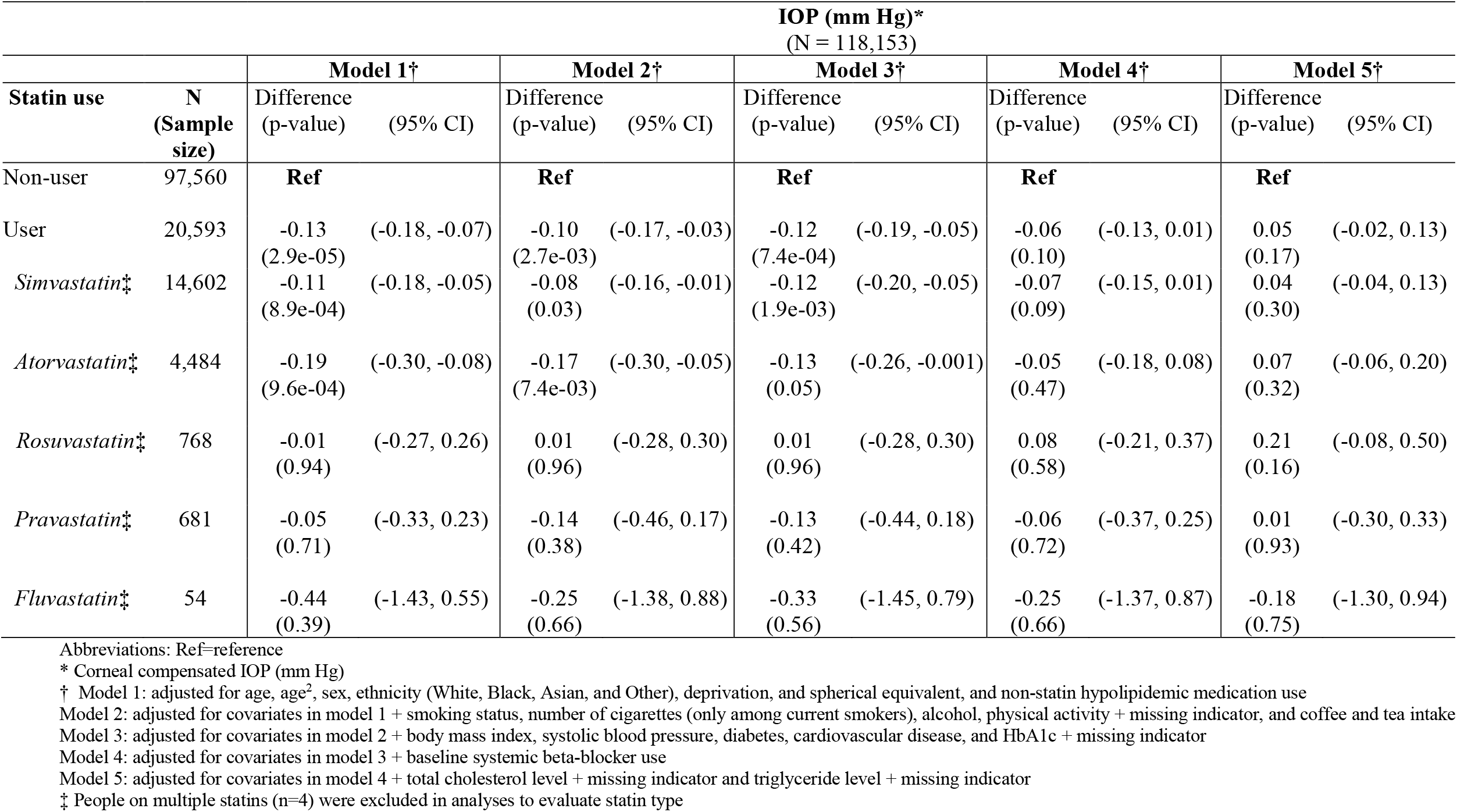
Association between statin use (2006-2010) and intraocular pressure (measured in 2009-2013) in the UK Biobank: multivariable- adjusted difference (95% CI) in IOP (mm Hg) by statin use

|  |  | IOP (mm Hg)*<br>(N = 118,153) |  |  |  |  |  |  |  |  |  |
| --- | --- | --- | --- | --- | --- | --- | --- | --- | --- | --- | --- |
| Statin use | N<br>(Sample size) | Model 1† |  | Model 2† |  | Model 3† |  | Model 4† |  | Model 5† |  |
|  |  | Difference<br>(p-value) | (95% CI) | Difference<br>(p-value) | (95% CI) | Difference<br>(p-value) | (95% CI) | Difference<br>(p-value) | (95% CI) | Difference<br>(p-value) | (95% CI) |
| Non-user | 97,560 | Ref |  | Ref |  | Ref |  | Ref |  | Ref |  |
| User | 20,593 | -0.13<br>(2.9e-05) | (-0.18, -0.07) | -0.10<br>(2.7e-03) | (-0.17, -0.03) | -0.12<br>(7.4e-04) | (-0.19, -0.05) | -0.06<br>(0.10) | (-0.13, 0.01) | 0.05<br>(0.17) | (-0.02, 0.13) |
| *Simvastatin*‡ | 14,602 | -0.11<br>(8.9e-04) | (-0.18, -0.05) | -0.08<br>(0.03) | (-0.16, -0.01) | -0.12<br>(1.9e-03) | (-0.20, -0.05) | -0.07<br>(0.09) | (-0.15, 0.01) | 0.04<br>(0.30) | (-0.04, 0.13) |
| *Atorvastatin*‡ | 4,484 | -0.19<br>(9.6e-04) | (-0.30, -0.08) | -0.17<br>(7.4e-03) | (-0.30, -0.05) | -0.13<br>(0.05) | (-0.26, -0.001) | -0.05<br>(0.47) | (-0.18, 0.08) | 0.07<br>(0.32) | (-0.06, 0.20) |
| *Rosuvastatin*‡ | 768 | -0.01<br>(0.94) | (-0.27, 0.26) | 0.01<br>(0.96) | (-0.28, 0.30) | 0.01<br>(0.96) | (-0.28, 0.30) | 0.08<br>(0.58) | (-0.21, 0.37) | 0.21<br>(0.16) | (-0.08, 0.50) |
| *Pravastatin*‡ | 681 | -0.05<br>(0.71) | (-0.33, 0.23) | -0.14<br>(0.38) | (-0.46, 0.17) | -0.13<br>(0.42) | (-0.44, 0.18) | -0.06<br>(0.72) | (-0.37, 0.25) | 0.01<br>(0.93) | (-0.30, 0.33) |
| *Fluvastatin*‡ | 54 | -0.44<br>(0.39) | (-1.43, 0.55) | -0.25<br>(0.66) | (-1.38, 0.88) | -0.33<br>(0.56) | (-1.45, 0.79) | -0.25<br>(0.66) | (-1.37, 0.87) | -0.18<br>(0.75) | (-1.30, 0.94) |

Abbreviations: Ref=reference
\* Corneal compensated IOP (mm Hg)
† Model 1: adjusted for age, age<sup>2</sup>, sex, ethnicity (White, Black, Asian, and Other), deprivation, and spherical equivalent, and non-statin hypolipidemic medication use
Model 2: adjusted for covariates in model 1 + smoking status, number of cigarettes (only among current smokers), alcohol, physical activity + missing indicator, and coffee and tea intake
Model 3: adjusted for covariates in model 2 + body mass index, systolic blood pressure, diabetes, cardiovascular disease, and HbA1c + missing indicator
Model 4: adjusted for covariates in model 3 + baseline systemic beta-blocker use
Model 5: adjusted for covariates in model 4 + total cholesterol level + missing indicator and triglyceride level + missing indicator
‡ People on multiple statins (n=4) were excluded in analyses to evaluate statin type

However, the relationship became nonsignificant and attenuated after additional adjustment for systemic beta-blocker use (Model 4: difference = -0.06 mm Hg; p=0.10) and remained nonsignificant in Model 5 that also controlled for serum total cholesterol level and triglyceride levels (Model 5: difference = 0.05 mm Hg; 95% CI: -0.02, 0.13; p=0.17). Similarly, in Model 5, specific statin types were not significantly associated with IOP (**Table 2**). In sensitivity analysis excluding prevalent glaucoma cases, the relation between statin use and IOP remained nonsignificant (Model 5: difference = 0.04 mm Hg; 95% CI: -0.04, 0.11; p=0.31; **Supplementary Table 3**). Results also remained unchanged in an alternative model where the reference group was restricted to participants not on any hypolipidemic agents (versus the larger group of non-statin users); furthermore, use of non-statin hypolipidemic drugs showed no association with IOP in a fully adjusted multivariable model (difference = 0.005 mm Hg; 95% CI: -0.14, 0.15; p=0.95; **Supplementary Table 4)**.

### Statin use and self-reported glaucoma in the UK Biobank

Among 192,283 participants including 8,982 cases of self-reported prevalent glaucoma, the relation between statin use and prevalent glaucoma was null in most models (Model 5: OR = 1.05; 95% CI: 0.98, 1.13; **Table 3**). When the reference group was restricted to participants not on any hypolipidemic treatments, the results were largely unchanged (**Supplementary Table 4**). Furthermore, non-statin hypolipidemic treatment was also not associated with prevalent glaucoma in a fully adjusted multivariable model (OR = 1.04; 95% CI: 0.92, 1.18; **Supplementary Table 4**).

**Table 3.**
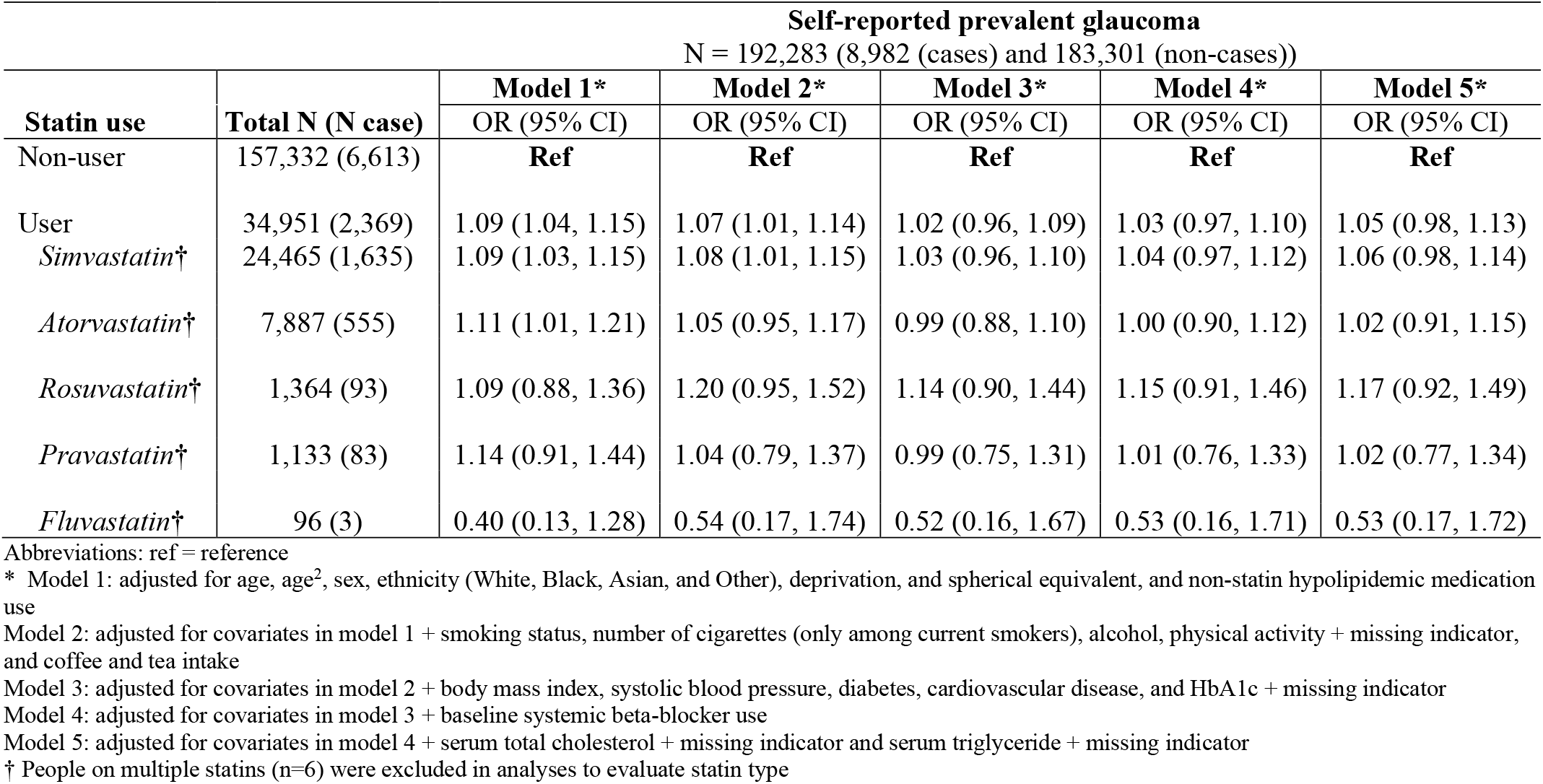
Association between statin use (2006-2010) and prevalent glaucoma in the UK Biobank (2006-2010): multivariable-adjusted odds ratio (95% CI)

### Statin use and glaucoma-related OCT parameters in the UK Biobank

The average thickness of mRNFL was 28.9 (3.8) microns and of mGCIPL was 75.2 (5.2) microns (**Supplementary Table 1**). In the basic multivariable model adjusted for age, age-squared, sex, ethnicity, deprivation, spherical equivalent and non-statin hypolipidemic treatments, statin use was associated with thinner mRNFL (Model 1: difference = -0.32 microns; p=8.6E-10) and mGCIPL (difference = -0.36 microns; p=1.3E-07, **Table 4**). In Model 5, the results for mRNFL were attenuated and only borderline significant (Model 5: difference = -0.14 microns; 95% CI: -0.28, -0.01; P=0.03). For mGCIPL, in model 5, results were attenuated, and non-significant (Model 5: difference = -0.12 microns; 95% CI: -0.29, 0.05; p=0.17). These results were nearly identical after excluding prevalent glaucoma cases (**Supplementary Table 3**).

**Table 4.**
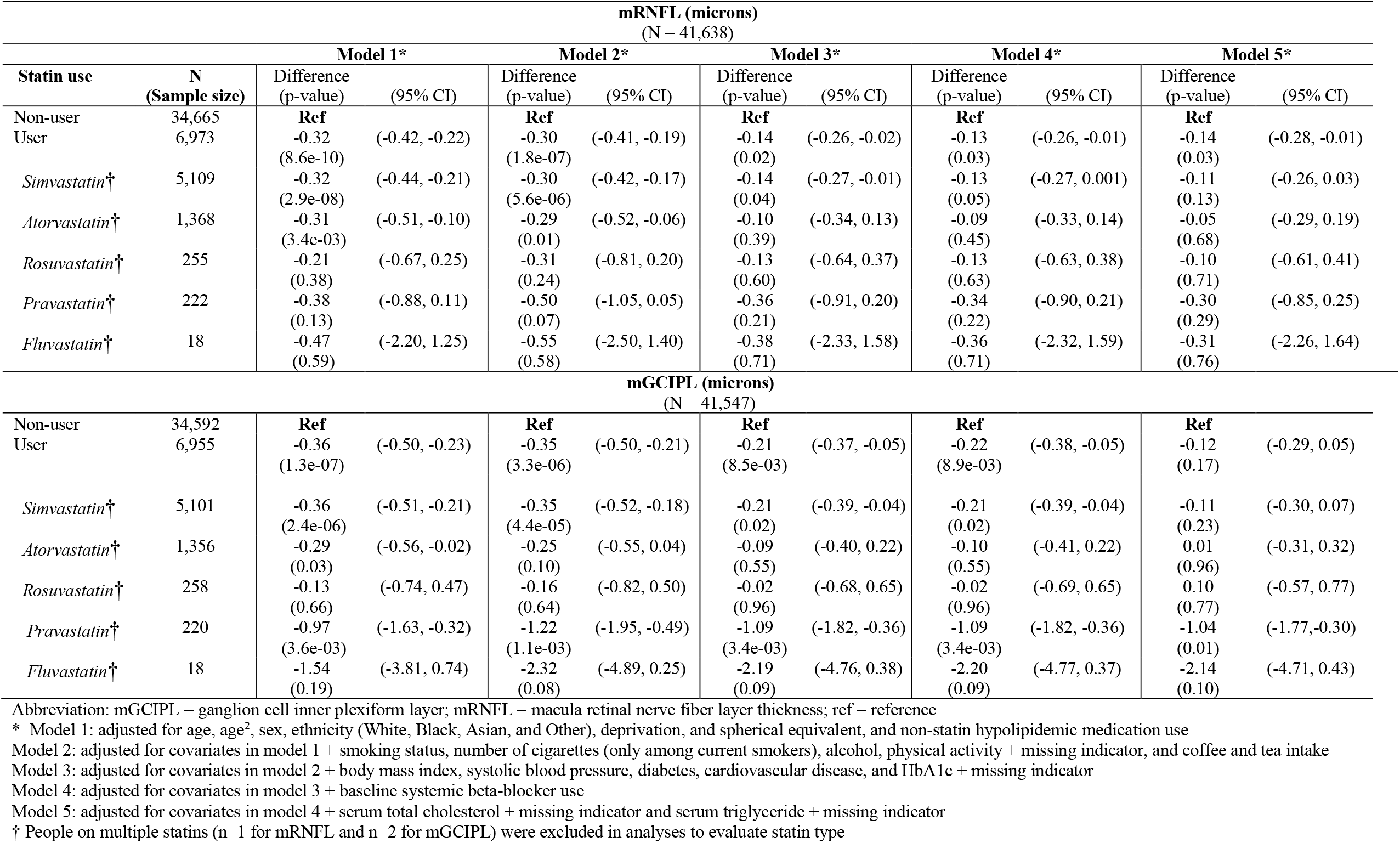
Association between statin use (2009-2010) and retinal nerve fiber layer (mRNFL) and ganglion cell plexiform layer (mGCIPL) thicknesses in the macula region (2009-2010) among UK Biobank participants

| mRNFL (microns)<br>(N = 41,638) |  |  |  |  |  |  |  |  |  |  |  |
| --- | --- | --- | --- | --- | --- | --- | --- | --- | --- | --- | --- |
|  |  | Model 1* |  | Model 2* |  | Model 3* |  | Model 4* |  | Model 5* |  |
| Statin use | N<br>(Sample size) | Difference<br>(p-value) | (95% CI) | Difference<br>(p-value) | (95% CI) | Difference<br>(p-value) | (95% CI) | Difference<br>(p-value) | (95% CI) | Difference<br>(p-value) | (95% CI) |
| Non-user | 34,665 | **Ref** |  | **Ref** |  | **Ref** |  | **Ref** |  | **Ref** |  |
| User | 6,973 | -0.32<br>(8.6e-10) | (-0.42, -0.22) | -0.30<br>(1.8e-07) | (-0.41, -0.19) | -0.14<br>(0.02) | (-0.26, -0.02) | -0.13<br>(0.03) | (-0.26, -0.01) | -0.14<br>(0.03) | (-0.28, -0.01) |
| *Simvastatin*† | 5,109 | -0.32<br>(2.9e-08) | (-0.44, -0.21) | -0.30<br>(5.6e-06) | (-0.42, -0.17) | -0.14<br>(0.04) | (-0.27, -0.01) | -0.13<br>(0.05) | (-0.27, 0.001) | -0.11<br>(0.13) | (-0.26, 0.03) |
| *Atorvastatin*† | 1,368 | -0.31<br>(3.4e-03) | (-0.51, -0.10) | -0.29<br>(0.01) | (-0.52, -0.06) | -0.10<br>(0.39) | (-0.34, 0.13) | -0.09<br>(0.45) | (-0.33, 0.14) | -0.05<br>(0.68) | (-0.29, 0.19) |
| *Rosuvastatin*† | 255 | -0.21<br>(0.38) | (-0.67, 0.25) | -0.31<br>(0.24) | (-0.81, 0.20) | -0.13<br>(0.60) | (-0.64, 0.37) | -0.13<br>(0.63) | (-0.63, 0.38) | -0.10<br>(0.71) | (-0.61, 0.41) |
| *Pravastatin*† | 222 | -0.38<br>(0.13) | (-0.88, 0.11) | -0.50<br>(0.07) | (-1.05, 0.05) | -0.36<br>(0.21) | (-0.91, 0.20) | -0.34<br>(0.22) | (-0.90, 0.21) | -0.30<br>(0.29) | (-0.85, 0.25) |
| *Fluvastatin*† | 18 | -0.47<br>(0.59) | (-2.20, 1.25) | -0.55<br>(0.58) | (-2.50, 1.40) | -0.38<br>(0.71) | (-2.33, 1.58) | -0.36<br>(0.71) | (-2.32, 1.59) | -0.31<br>(0.76) | (-2.26, 1.64) |
| mGCIPL (microns)<br>(N = 41,547) |  |  |  |  |  |  |  |  |  |  |  |
| Non-user | 34,592 | **Ref** |  | **Ref** |  | **Ref** |  | **Ref** |  | **Ref** |  |
| User | 6,955 | -0.36<br>(1.3e-07) | (-0.50, -0.23) | -0.35<br>(3.3e-06) | (-0.50, -0.21) | -0.21<br>(8.5e-03) | (-0.37, -0.05) | -0.22<br>(8.9e-03) | (-0.38, -0.05) | -0.12<br>(0.17) | (-0.29, 0.05) |
| *Simvastatin*† | 5,101 | -0.36<br>(2.4e-06) | (-0.51, -0.21) | -0.35<br>(4.4e-05) | (-0.52, -0.18) | -0.21<br>(0.02) | (-0.39, -0.04) | -0.21<br>(0.02) | (-0.39, -0.04) | -0.11<br>(0.23) | (-0.30, 0.07) |
| *Atorvastatin*† | 1,356 | -0.29<br>(0.03) | (-0.56, -0.02) | -0.25<br>(0.10) | (-0.55, 0.04) | -0.09<br>(0.55) | (-0.40, 0.22) | -0.10<br>(0.55) | (-0.41, 0.22) | 0.01<br>(0.96) | (-0.31, 0.32) |
| *Rosuvastatin*† | 258 | -0.13<br>(0.66) | (-0.74, 0.47) | -0.16<br>(0.64) | (-0.82, 0.50) | -0.02<br>(0.96) | (-0.68, 0.65) | -0.02<br>(0.96) | (-0.69, 0.65) | 0.10<br>(0.77) | (-0.57, 0.77) |
| *Pravastatin*† | 220 | -0.97<br>(3.6e-03) | (-1.63, -0.32) | -1.22<br>(1.1e-03) | (-1.95, -0.49) | -1.09<br>(3.4e-03) | (-1.82, -0.36) | -1.09<br>(3.4e-03) | (-1.82, -0.36) | -1.04<br>(0.01) | (-1.77, -0.30) |
| *Fluvastatin*† | 18 | -1.54<br>(0.19) | (-3.81, 0.74) | -2.32<br>(0.08) | (-4.89, 0.25) | -2.19<br>(0.09) | (-4.76, 0.38) | -2.20<br>(0.09) | (-4.77, 0.37) | -2.14<br>(0.10) | (-4.71, 0.43) |

Abbreviation: mGCIPL = ganglion cell inner plexiform layer; mRNFL = macula retinal nerve fiber layer thickness; ref = reference
\* Model 1: adjusted for age, age<sup>2</sup>, sex, ethnicity (White, Black, Asian, and Other), deprivation, and spherical equivalent, and non-statin hypolipidemic medication use
Model 2: adjusted for covariates in model 1 + smoking status, number of cigarettes (only among current smokers), alcohol, physical activity + missing indicator, and coffee and tea intake
Model 3: adjusted for covariates in model 2 + body mass index, systolic blood pressure, diabetes, cardiovascular disease, and HbA1c + missing indicator
Model 4: adjusted for covariates in model 3 + baseline systemic beta-blocker use
Model 5: adjusted for covariates in model 4 + serum total cholesterol + missing indicator and serum triglyceride + missing indicator
† People on multiple statins (n=1 for mRNFL and n=2 for mGCIPL) were excluded in analyses to evaluate statin type

Furthermore, in models where the reference group was restricted to participants who did not take any hypolipidemic treatments, the results were very similar (**Supplementary Table 4)**.

### Genetic modification of statin use – glaucoma-related outcomes

To determine whether the relation between statin use and glaucoma-related outcomes may differ by genetic propensity for glaucoma, we evaluated whether the MTAG PRS * statin interaction term was significant when added to Model 5. The glaucoma MTAG PRS did not significantly modify the relation between statin use and IOP, prevalent glaucoma, mRNFL thickness or mGCIPL thickness (P_interaction_≥0.16; **Table 5**).

**Table 5.** Multivariable-adjusted interactions between statin use and the glaucoma MTAG GRS on four glaucoma-related outcomes in the UK Biobank*

|  | **IOP (mmHg)** | **Prevalent glaucoma** | **mRNFL (microns)** | **mGCIPL (microns)** |
| --- | --- | --- | --- | --- |
|  | (N = 118,153) | (N = 192,283) | (N = 41,638) | (N = 41,547) |
|  | Beta<sub>int</sub> (P<sub>int</sub>) | OR<sub>int</sub> (P<sub>int</sub>) | Beta<sub>int</sub> (P<sub>int</sub>) | Beta<sub>int</sub> (P<sub>int</sub>) |
| **MTAG GRS\*Statin use**<br>(GRS*statin use) | 0.05 (0.16) | 0.99 (0.72) | 0.04 (0.51) | -0.09 (0.27) |

Abbreviations: GRS = genetic risk score, int = interaction; OR = Odds Ratio; MTAG = multi-trait association of genome wide association studies
\* Model for IOP: adjusted for age, age<sup>2</sup>, sex, ethnicity (White, Black, Asian, and Other), deprivation, spherical equivalent, smoking status, number of cigarettes (only among current smokers), alcohol, physical activity + missing indicator, coffee and tea intake, body mass index, systolic blood pressure, diabetes, cardiovascular disease (CVD), HbA1C + missing indicator, systemic beta-blocker use, serum total cholesterol (TC), serum triglycerides (TG) + missing indicator + non-statin hypolipidemic drug use + TC\*MTAG GRS + CVD\*MTAG GRS + TG\*MTAG GRS + age\*MTAG GRS + age<sup>2</sup>\*MTAG GRS + non-statin hypolipidemic drug use\*MTAG GRS
Model for prevalent glaucoma: adjusted for age, age<sup>2</sup>, sex, ethnicity (White, Black, Asian, and Other), deprivation, spherical equivalent + missing indicator, smoking status, number of cigarettes (only among current smokers), alcohol, physical activity + missing indicator, coffee and tea intake, body mass index, systolic blood pressure, diabetes, cardiovascular disease (CVD), HbA1C, systemic beta-blocker use, serum total cholesterol (TC), serum triglycerides (TG) + non-statin hypolipidemic drug use + TC\*MTAG GRS + CVD\*MTAG GRS + TG\*MTAG GRS + age\*MTAG GRS + age<sup>2</sup>\*MTAG GRS + non-statin hypolipidemic drug use\*MTAG GRS
Model for RNFL and GCIPL: adjusted for age, age<sup>2</sup>, sex, ethnicity (White, Black, Asian, and Other), deprivation, spherical equivalent, smoking status, number of cigarettes (only among current smokers), alcohol, physical activity + missing indicator, coffee and tea intake, body mass index, systolic blood pressure, diabetes, cardiovascular disease (CVD), HbA1C + missing indicator, systemic beta-blocker use, serum total cholesterol (TC), serum triglycerides (TG) + missing indicator + non-statin hypolipidemic drug use + TC\*MTAG GRS + CVD\*MTAG GRS + TG\*MTAG GRS + age\*MTAG GRS + age<sup>2</sup>\*MTAG GRS + non-statin hypolipidemic drug use\*MTAG GRS

Furthermore, the glaucoma MTAG PRS did not significantly modify the relation between non-statin hypolipidemic medicine use and the glaucoma-related outcomes (P_interaction_ ≥ 0.24; **Supplementary Table 5)**.

### Mendelian Randomization (MR) Analyses

MR analyses showed no evidence of a causal link between HMG-CoA reductase activity and any of POAG, VCDR, IOP, mRNFL or mGCIPL (**Supplementary Table 6** and **Supplementary Figure 1**). Only the analysis using the POAG dataset showed significant unbalanced horizontal pleiotropy as well as global heterogeneity (Cochran’s Q p=0.05, *I*^2^ =57.2).

**Supplementary Figure 2** shows power calculations and highlights the minimum ORs and differences (betas) to achieve 80% power for each outcome. We compared these to the published estimates of effect sizes where available, as well as our effect estimates. These calculations showed that while we were well powered to detect a significant effect in the VCDR outcome, we were less well powered for the other outcomes.

### Other secondary analyses

Among UK Biobank participants with total serum cholesterol <200 mg/d (5.13mmol/L) and no history of cardiovascular disease, diabetes or systemic beta blocker use, there was also no relationship between statin use and IOP, prevalent glaucoma or mRNFL in multivariable analysis (p≥0.17; **Supplementary Table 7**), while statin use was associated with thinner mGCIPL thickness (difference = -0.42 microns; p=0.01; **Supplementary Table 7)**. Finally, in a multivariable analysis of statin use in relation to the glaucoma outcomes among people with complete data on all glaucoma-related parameters (N=41,444), the same general trends were observed as those in the main analyses (**Supplementary Table 8)**.

## Discussion

In this large study in the UK Biobank, based on the main analyses as well as the MR analysis, we did not observe any beneficial associations of statin use in relation to glaucoma prevalence or glaucoma-related traits (IOP, mRNFL or mGCIPL); furthermore, the null associations were not modified by genetic predisposition to glaucoma.

For IOP, our nested models revealed that the relation between statin use and IOP was subject to considerable confounding, but ultimately, in a fully adjusted model, the relationship was null (n=118,153). Similarly, we did not observe that non-statin hypolipidemic treatment was associated with IOP. While one Singaporean study (n=10,033) observed that statin use was associated with higher IOP,^26^ our null findings are consistent with other studies that have observed null associations with IOP.^6, 27^ For example, a Canadian pharmaceutical database study (n=8548) showed no difference in adjunct topical IOP-lowering medication use between statin users and nonusers.^28^ In our study, one of the strongest confounders for this association was systemic beta-blocker use. This was consistent with results from another British study of 7093 participants showing that the relation between statin use and IOP was no longer significant after controlling for systemic beta-blocker use.^29^ Another confounder was total cholesterol; indeed, a recent meta-analysis suggested that total cholesterol is a risk factor for higher IOP.^30^ In our nested models, we were able to adjust for systemic beta-blocker use, total cholesterol levels and other conditions that may be indications for statins.

The relation between statin use and glaucoma has also been intensely studied and has yielded very mixed results ranging from inverse,^6, 31-34^ to null,^7, 35-37^ to adverse.^8, 38^ Different ways of ascertaining statin use, dissimilar definitions of glaucoma, varied study methodologies, and uneven follow-up periods may have played a role in the inconsistent results. Furthermore, it is difficult to address residual confounding and to disentangle the effects of statins from the underlying indications for statins, such as dyslipidemia, cardiovascular disease, and diabetes. No observational study on this subject is ideal, and our analysis lacked information on dosage or duration of statin use and was a cross-sectional study that assessed glaucoma by self-report. However, our cross-sectional study results were consistent with a meta-analysis that observed that use of statins for 2 or more years was not associated with glaucoma incidence.^6^ Also, our study had a large sample size, was able to control for multiple indications for statins including total serum cholesterol, evaluated multiple glaucoma outcomes and the main analyses were augmented with an MR study, which provide a way to evaluate dose-response relationships as well as relationships with life-time exposure.^9^

Our study was the first to assess the relation between statin use and inner retinal structure. Because early glaucoma can affect the structures in the macula region,^39^ OCT scanning of the macula region can be useful in diagnosing glaucoma;^18, 19^ indeed, a previous UK Biobank study reported that higher IOP was associated with a thinner mGCIPL (but there was no relation with mRNFL thickness).^17^ However, we observed no beneficial associations between statin use and macular OCT structural parameters (mRNFL and mGCIPL); an initial significant adverse association with thinner measures was attenuated after adjustment for BMI, systolic blood pressure, diabetes, and cardiovascular disease, although for mRNFL, the adverse association remained nominally significant. The findings with OCT structural parameters collectively support the notion that statin use is not associated with glaucoma.

We also assessed whether a glaucoma MTAG PRS might interact with statin use to influence various glaucoma-related outcomes, but we did not observe any significant interactions. It is important to keep in mind that this same MTAG PRS was a powerful tool that predicted earlier age at glaucoma diagnosis, glaucoma disease progression and the need for glaucoma surgery.^21^ Furthermore, in MR analyses, we used a genetic instrument to approximate HMG-CoA activity, an instrument that has been shown to predict cardiovascular disease.^10^ Although we had only modest power to find associations in MR analyses, we were able to preclude strong associations between statin use with various glaucoma outcomes, further supporting the lack of a strong association of statin use on glaucoma. Finally, we cannot be certain that the HMG-CoA reductase variants operate directly and solely through the HMG-CoA reductase gene, although there is some evidence that these variants alter splicing of HMG-CoA reductase transcripts.^10^

Our study had several limitations. This was a cross-sectional study that evaluated prevalent glaucoma and IOP and OCT measures obtained at a single timepoint. This makes causal inferences quite challenging; thus, our results should be interpreted with caution. Also, the definition of glaucoma was not highly specific and relied on participant reporting, so our results may have been susceptible to various biases related to misclassification of the outcome. While we had data on the type of statin use as well as data on non-statin hypolipidemic medicines, we did not have data on dosage or duration of statin use; this was a limitation as studies have found relationships with longer duration of use, although the MR analyses provided some complementary data representing dose-response relations and life-time exposures. Finally, because <8% of our study population was nonwhite, our findings may not be generalizable to people of color.

This study had several strengths. This was a large study with multiple glaucoma traits, covariates, and genotype information. Current statin use and type of statin information was collected with the help of an in-person interviewer, and as expected, those on statins had lower serum total cholesterol than nonusers, providing a measure of construct validity for this exposure. We also generated a genetic instrument that mimicked variation in HMG-CoA reductase activity and served as a surrogate of life-time statin use. This was also the first study to assess whether a glaucoma MTAG PRS might modify the relationship between statin use and glaucoma outcomes.

Overall, our study provides additional evidence that statin use is not favorably associated with glaucoma-related outcomes and that statin use may not be an effective primary glaucoma prevention strategy. Future large prospective studies with adequate control for major confounders, including other medications and indications for statin use, duration information as well as objective measures such as peripapillary RNFL thickness and a focus on ethnic minorities could shed more light on this subject.

## Data Availability

All data are available at ukbiobank.ac.uk.

## Abbreviations and Acronyms

BMI: body mass index
CI: confidence interval
GWAS: genome-wide association study
HMG-CoA: 3-hydroxy-3-methylglutaryl-coenzyme A reductase
IOP: intraocular pressure
IOPcc: corneal-compensated IOP
LDL: low density lipoprotein
MET: metabolic equivalent of task
mGCIPL: macular ganglion cell inner plexiform layer
MR: Mendelian Randomization
mRNFL: macular retinal nerve fiber layer
MTAG: multi-trait analysis of genome wide association study
OCT: ocular coherence tomography
OR: odds ratio
POAG: primary open-angle glaucoma
PRS: polygenic risk score
SD: standard deviation
SNP: single nucleotide polymorphism
VCDR: vertical cup disc ratio.

## Notes

Financial Support: NEI R01 EY015473; NEI R01 EY032559; NEI R01 EY022305 and unrestricted Challenge Grant from Research to Prevent Blindness (NYC) and The Glaucoma Foundation (NYC). APK was supported by a UK Research and Innovation Future Leaders Fellowship and an Alcon Research Institute Young Investigator Award. Dr. Ron Do is supported by the National Institute of General Medical Sciences of the NIH (R35 GM124836) and the National Heart, Lung, and Blood Institute of the NIH (R01 HL139865 and R01 HL155915). The sponsor or funding organization had no role in the design or conduct of this research.

Conflict of Interest: No conflicting relationship exists for authors except for the following authors: **LRP:** Consultant to Twenty Twenty, Skye Biosciences, and Eyenovia. **APK:** Consultant to Abbvie, Google Health, Reichert, Santen; Lecture fees from Thea. **RD:** Received grants from AstraZeneca, grants, and non-financial support from Goldfinch Bio; Scientific co-founder, consultant and equity holder for Pensieve Health, and Consultant for Variant Bio. **JLW:** Consultant to Allergan, Editas, Broadwing Bio, Maze, Regenxbio, Aerpio; Research support from Aerpio.

### Competing Interest Statement

LRP: Consultant to Twenty Twenty, Skye Biosciences, and Eyenovia. APK: Consultant to Abbvie, Google Health, Reichert, Santen; Lecture fees from Thea. RD: Received grants from AstraZeneca, grants, and non-financial support from Goldfinch Bio; Scientific co-founder, consultant and equity holder for Pensieve Health, and Consultant for Variant Bio. JLW: Consultant to Allergan, Editas, Broadwing Bio, Maze, Regenxbio, Aerpio; Research support from Aerpio.

### Funding Statement

NEI R01 EY015473; NEI R01 EY032559; NEI R01 EY022305 and unrestricted Challenge Grant from Research to Prevent Blindness (NYC) and The Glaucoma Foundation (NYC). APK was supported by a UK Research and Innovation Future Leaders Fellowship and an Alcon Research Institute Young Investigator Award. Dr. Ron Do is supported by the National Institute of General Medical Sciences of the NIH (R35 GM124836) and the National Heart, Lung, and Blood Institute of the NIH (R01 HL139865 and R01 HL155915). The sponsor or funding organization had no role in the design or conduct of this research.

### Author Declarations

Data for this study was provided by the UK Biobank under Application Number 36741.

## References

1. Salami JA, Warraich H, Valero-Elizondo J, et al. National Trends in Statin Use and Expenditures in the US Adult Population From 2002 to 2013: Insights From the Medical Expenditure Panel Survey. JAMA Cardiol. 2017;2:56–65.

2. US Preventive Services Task Force, Bibbins-Domingo K, Grossman DC, et al. Statin Use for the Primary Prevention of Cardiovascular Disease in Adults: US Preventive Services Task Force Recommendation Statement. JAMA. 2016;316:1997–2007.

3. O’Keeffe AG, Nazareth I, Petersen I. Time trends in the prescription of statins for the primary prevention of cardiovascular disease in the United Kingdom: a cohort study using The Health Improvement Network primary care data. Clin Epidemiol. 2016;8:123–132.

4. Pranata R, Vania R, Victor AA. Statin reduces the incidence of diabetic retinopathy and its need for intervention: A systematic review and meta-analysis. Eur J Ophthalmol. 2021;31:1216–1224.

5. Ludwig CA, Vail D, Rajeshuni NA, et al. Statins and the progression of age-related macular degeneration in the United States. PLoS One. 2021;16:e0252878.

6. McCann P, Hogg RE, Fallis R, Azuara-Blanco A. The Effect of Statins on Intraocular Pressure and on the Incidence and Progression of Glaucoma: A Systematic Review and Meta-Analysis. Invest Ophthalmol Vis Sci. 2016;57:2729–2748.

7. Kang JH, Boumenna T, Stein JD, et al. Notice of Retraction and Replacement. Kang et al. Association of statin use and high serum cholesterol levels with risk of primary open-angle glaucoma. JAMA Ophthalmol. 2019;137(7):756-765. JAMA Ophthalmol. 2020;138:588–589.

8. Yuan Y, Wang W, Shang X, et al. Association between statin use and the risks of glaucoma in Australia: a 10-year cohort study. Br J Ophthalmol. 2021.

9. Davey Smith G, Hemani G. Mendelian randomization: genetic anchors for causal inference in epidemiological studies. Hum Mol Genet. 2014;23:R89–98.

10. Ference BA, Robinson JG, Brook RD, et al. Variation in PCSK9 and HMGCR and Risk of Cardiovascular Disease and Diabetes. N Engl J Med. 2016;375:2144–2153.

11. Willer CJ, Schmidt EM, Sengupta S, et al. Discovery and refinement of loci associated with lipid levels. Nat Genet. 2013;45:1274–1283.

12. Chua SYL, Thomas D, Allen N, et al. Cohort profile: design and methods in the eye and vision consortium of UK Biobank. BMJ Open. 2019;9:e025077.

13. Luce DA. Determining in vivo biomechanical properties of the cornea with an ocular response analyzer. J Cataract Refract Surg. 2005;31:156–162.

14. Khawaja AP, Cooke Bailey JN, Wareham NJ, et al. Genome-wide analyses identify 68 new loci associated with intraocular pressure and improve risk prediction for primary open-angle glaucoma. Nat Genet. 2018;50:778–782.

15. MacGregor S, Ong JS, An J, et al. Genome-wide association study of intraocular pressure uncovers new pathways to glaucoma. Nat Genet. 2018;50:1067–1071.

16. Hysi PG, Cheng CY, Springelkamp H, et al. Genome-wide analysis of multi-ancestry cohorts identifies new loci influencing intraocular pressure and susceptibility to glaucoma. Nat Genet. 2014;46:1126–1130.

17. Khawaja AP, Chua S, Hysi PG, et al. Comparison of Associations with Different Macular Inner Retinal Thickness Parameters in a Large Cohort: The UK Biobank. Ophthalmology. 2020;127:62–71.

18. Kim KE, Park KH. Macular imaging by optical coherence tomography in the diagnosis and management of glaucoma. Br J Ophthalmol. 2018;102:718–724.

19. Oddone F, Lucenteforte E, Michelessi M, et al. Macular versus Retinal Nerve Fiber Layer Parameters for Diagnosing Manifest Glaucoma: A Systematic Review of Diagnostic Accuracy Studies. Ophthalmology. 2016;123:939–949.

20. Wain LV, Shrine N, Miller S, et al. Novel insights into the genetics of smoking behaviour, lung function, and chronic obstructive pulmonary disease (UK BiLEVE): a genetic association study in UK Biobank. Lancet Respir Med. 2015;3:769–781.

21. Craig JE, Han X, Qassim A, et al. Multitrait analysis of glaucoma identifies new risk loci and enables polygenic prediction of disease susceptibility and progression. Nat Genet. 2020;52:160–166.

22. Gharahkhani P, Jorgenson E, Hysi P, et al. Genome-wide meta-analysis identifies 127 open-angle glaucoma loci with consistent effect across ancestries. Nat Commun. 2021;12:1258.

23. Springelkamp H, Iglesias AI, Mishra A, et al. New insights into the genetics of primary open-angle glaucoma based on meta-analyses of intraocular pressure and optic disc characteristics. Hum Mol Genet. 2017;26:438–453.

24. Currant H, Hysi P, Fitzgerald TW, et al. Genetic variation affects morphological retinal phenotypes extracted from UK Biobank optical coherence tomography images. PLoS Genet. 2021;17:e1009497.

25. Keller MC. Gene x environment interaction studies have not properly controlled for potential confounders: the problem and the (simple) solution. Biol Psychiatry. 2014;75:18–24.

26. Ho H, Shi Y, Chua J, et al. Association of Systemic Medication Use With Intraocular Pressure in a Multiethnic Asian Population: The Singapore Epidemiology of Eye Diseases Study. JAMA Ophthalmol. 2017;135:196–202.

27. Hohn R, Mirshahi A, Nickels S, et al. Cardiovascular medication and intraocular pressure: results from the Gutenberg Health Study. Br J Ophthalmol. 2017;101:1633–1637.

28. Iskedjian M, Walker JH, Desjardins O, et al. Effect of selected antihypertensives, antidiabetics, statins and diuretics on adjunctive medical treatment of glaucoma: a population based study. Curr Med Res Opin. 2009;25:1879–1888.

29. Khawaja AP, Chan MP, Broadway DC, et al. Systemic medication and intraocular pressure in a British population: the EPIC-Norfolk Eye Study. Ophthalmology. 2014;121:1501–1507.

30. Wang S, Bao X. Hyperlipidemia, Blood Lipid Level, and the Risk of Glaucoma: A Meta-Analysis. Invest Ophthalmol Vis Sci. 2019;60:1028–1043.

31. Marcus MW, Muskens RP, Ramdas WD, et al. Cholesterol-lowering drugs and incident open-angle glaucoma: a population-based cohort study. PLoS One. 2012;7:e29724.

32. McGwin G, Jr., McNeal S, Owsley C, et al. Statins and other cholesterol-lowering medications and the presence of glaucoma. Arch Ophthalmol. 2004;122:822–826.

33. Stein JD, Newman-Casey PA, Talwar N, et al. The relationship between statin use and open-angle glaucoma. Ophthalmology. 2012;119:2074–2081.

34. Talwar N, Musch DC, Stein JD. Association of Daily Dosage and Type of Statin Agent With Risk of Open-Angle Glaucoma. JAMA Ophthalmol. 2017;135:263–267.

35. Owen CG, Carey IM, Shah S, et al. Hypotensive medication, statins, and the risk of glaucoma. Invest Ophthalmol Vis Sci. 2010;51:3524–3530.

36. Zheng W, Dryja TP, Wei Z, et al. Systemic Medication Associations with Presumed Advanced or Uncontrolled Primary Open-Angle Glaucoma. Ophthalmology. 2018;125:984–993.

37. Shon K, Sung KR. Dyslipidemia, Dyslipidemia Treatment, and Open-angle Glaucoma in the Korean National Health and Nutrition Examination Survey. J Glaucoma. 2019;28:550–556.

38. Chen HY, Hsu SY, Chang YC, et al. Association Between Statin Use and Open-angle Glaucoma in Hyperlipidemia Patients: A Taiwanese Population-based Case-control Study. Medicine (Baltimore). 2015;94:e2018.

39. Hood DC, Raza AS, de Moraes CG, et al. Glaucomatous damage of the macula. Prog Retin Eye Res. 2013;32:1–21.

