## Supplementary Methods for "Statin use in relation to intraocular pressure, glaucoma, and ocular coherence tomography parameters in the UK Biobank"

### Supplementary Methods for Mendelian Randomization Experiments

Mendelian Randomization (MR) is the use of genetic variants as proxies for modifiable risk factors and therapeutics in a way that has been likened to a randomized control trial by genetic proxy. The fixed nature of genetic variants reduces bias from environment exposure and reverse causation.

As statin's cholesterol reduction and pleiotropic effects appear mediated by inhibition of 3-hydroxy-3-methylglutaryl-coenzyme A (HMG-CoA) reductase, it would be expected that if statins protect against primary open-angle glaucoma (POAG), subjects with genetically modified activity of this enzyme from birth should have a commensurate difference in risk of POAG. We conducted a MR study using gene variants of HMG-CoA reductase to see whether these have any bearing on the genetic risk of primary open angle glaucoma (POAG),[1] intraocular pressure (IOP),[2] vertical cup-disc ratio (VCDR),[3] macular retinal nerve fiber layer (mRNFL) thickness and the macular ganglion cell inner plexiform layer (mGCIPL) thickness.[4]

We performed the inverse-variance weighted (IVW) analyses accounting for correlations between the variants[5, 6] using a random-effects model. Sensitivity analyses included weighted median and egger regression. Power calculations were performed to estimate the minimum odds ratio or beta required for 80% power.[7] Global heterogeneity was measured using Cochran's Q and $I^2$ statistic. All analyses were performed using R (version 1.4.17) using R packages "TwosampleMR"[8, 9] and "MendelianRandomization".

Variants in HMG-CoA reductase were obtained from the Global Lipids Genetic Consortium (GLGC).[10] We used serum LDL as a biomarker for the effect size and scaled to one standard deviation increase in LDL-cholesterol (39mg/dl). The GLGC involved up to 188,577 individuals of European ancestry who were not on any lipid-lowering medication. Six variants that are conditionally associated with LDL and are not strongly correlated ($r^2$ <0.4) were originally chosen by Ference et al.[11] Together, they account for 0.4% of the variance in circulating LDL. For our outcome datasets, we selected the largest published GWAS available performed in a European population and full details can be found in the source paper: POAG (*n*=216,257) ,[1] IOP (*n*=139,555),[2] VCDR (*n*=23,899),[3] mRNFL (*n*=31,434)[4] and mGCIPL (*n*=31,434).[4]

For the POAG dataset, we used the stage 1 genome-wide association study (GWAS) summary data from a recent meta-analysis that combined 18 POAG studies using ICD9/10 coding data in Europeans (16,677 POAG cases vs 199,580 controls).[1]

The IOP GWAS[2] is the largest performed to date, including 139,555 participants from three cohorts: UK Biobank,[12] EPIC-Norfolk[13] and the previously reported combined results from 14 European studies in the International Glaucoma Genetics Consortium (IGGC).[3]

The VCDR GWAS was performed as part of the metanalysis by the IGGC, combining multiple studies and included *n* = 23,899 participants of European descent.[3]

The mRNFL and mGCIPL GWAS used data solely from UK Biobank. Ocular coherence tomography (OCT) macula scans that fit quality control measures were extracted from the densest populated well-mixed population within the overall dataset (n= 31,434).[4]

### REFERENCES

1. Gharahkhani P, Jorgenson E, Hysi P, et al. Genome-wide meta-analysis identifies 127 open-angle glaucoma loci with consistent effect across ancestries. *Nat Commun.* 2021;12:1258.
2. Khawaja AP, Cooke Bailey JN, Wareham NJ, et al. Genome-wide analyses identify 68 new loci associated with intraocular pressure and improve risk prediction for primary open-angle glaucoma. *Nat Genet.* 2018;50:778-782.
3. Springelkamp H, Iglesias AI, Mishra A, et al. New insights into the genetics of primary open-angle glaucoma based on meta-analyses of intraocular pressure and optic disc characteristics. *Hum Mol Genet.* 2017;26:438-453.
4. Currant H, Hysi P, Fitzgerald TW, et al. Genetic variation affects morphological retinal phenotypes extracted from UK Biobank optical coherence tomography images. *PLoS Genet.* 2021;17:e1009497.
5. Burgess S, Dudbridge F, Thompson SG. Combining information on multiple instrumental variables in Mendelian randomization: Comparison of allele score and summarized data methods. *Stat Med.* 2016;35:1880-1906.
6. Yavorska OO, Burgess S. MendelianRandomization: An R package for performing Mendelian randomization analyses using summarized data. *Int J Epidemiol.* 2017;46:1734-1739.
7. Brion MJ, Shakhbazov K, Visscher PM. Calculating statistical power in Mendelian randomization studies. *Int J Epidemiol.* 2013;42:1497-1501.
8. Hemani G, Zheng J, Elsworth B, et al. The MR-base platform supports systematic causal inference across the human phenome. *Elife.* 2018;7:e34408
9. Hemani G, Tilling K, Davey Smith G. Orienting the causal relationship between imprecisely measured traits using GWAS summary data. *PLoS Genet.* 2017;13:e1007081.
10. Willer CJ, Schmidt EM, Sengupta S, et al. Discovery and refinement of loci associated with lipid levels. *Nat Genet.* 2013;45:1274-1283.
11. Ference BA, Robinson JG, Brook RD, et al. Variation in PCSK9 and HMGCR and risk of cardiovascular disease and diabetes. *N Engl J Med.* 2016;375:2144-2153.
12. Chan MP, Grossi CM, Khawaja AP, et al. Associations with intraocular pressure in a large cohort: Results from the UK Biobank. *Ophthalmology.* 2016;123:771-782.
13. Khawaja AP, Chan MP, Hayat S, et al. The EPIC-Norfolk eye study: Rationale, methods and a cross-sectional analysis of visual impairment in a population-based cohort. *BMJ Open.* 2013;3:e002684.
