## Supplementary Figure 1 for "Statin use in relation to intraocular pressure, glaucoma, and ocular coherence tomography parameters in the UK Biobank"

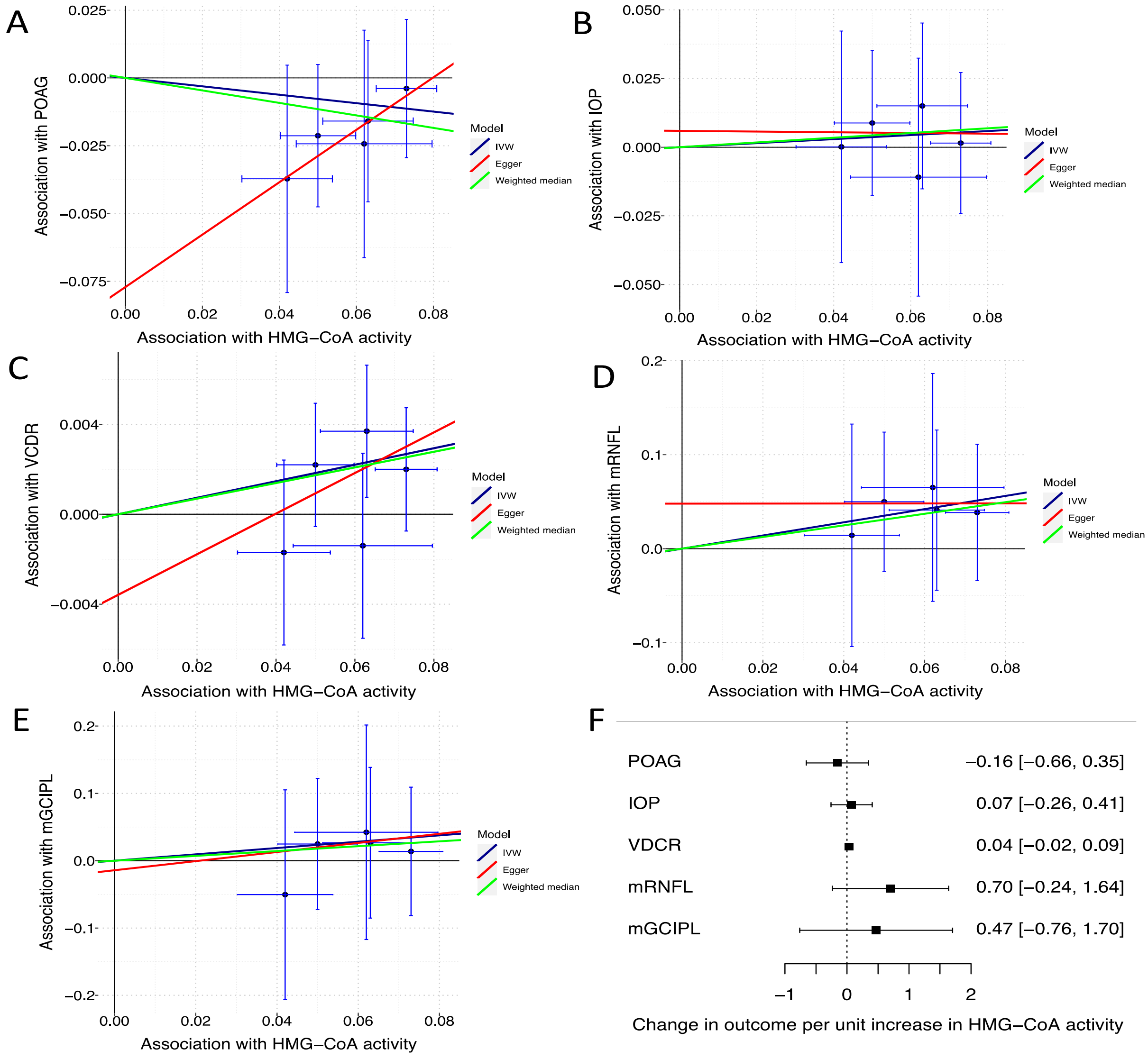


### Supplementary Figure 1.

Panels A-E show scatter plots of the association between 3-hydroxy-3-methylglutaryl-coenzyme A (HMG-CoA) reductase activity (measured in standard deviation LDL increase, where 1 standard deviation is 39mg/dl) and outcomes using three sensitivity analyses; HMG-CoA reductase variant rs2006760 was removed prior to analysis for being palindromic with intermediate allele frequencies, leaving five variants. Panel F shows the forest plot of the effect of change per unit increase in HMG-CoA reductase activity (standard deviation increase in LDL) on each outcome; estimates represent the Inverse Variance Weighted (IVW) analysis with 95% confidence intervals. The analysis using the POAG dataset showed significant unbalanced horizontal pleiotropy as well as global heterogeneity (Cohran's Q p=0.05, I2 =57.2); however, all other analyses did not. Overall, the IVW analyses showed no evidence of a causal link between HMG-CoA reductase activity and any of POAG, VCDR, IOP, mRNFL or mGCIPL

Abbreviations: IOP=intraocular pressure; mGCIPL= macular ganglion cell inner plexiform layer thickness in the macular region; mRNFL= retinal nerve fiber layer thickness in the macular region; POAG = primary open angle glaucoma; VCDR = vertical cup disc ratio.
