## Supplementary Figure 2 for "Statin use in relation to intraocular pressure, glaucoma, and ocular coherence tomography parameters in the UK Biobank"

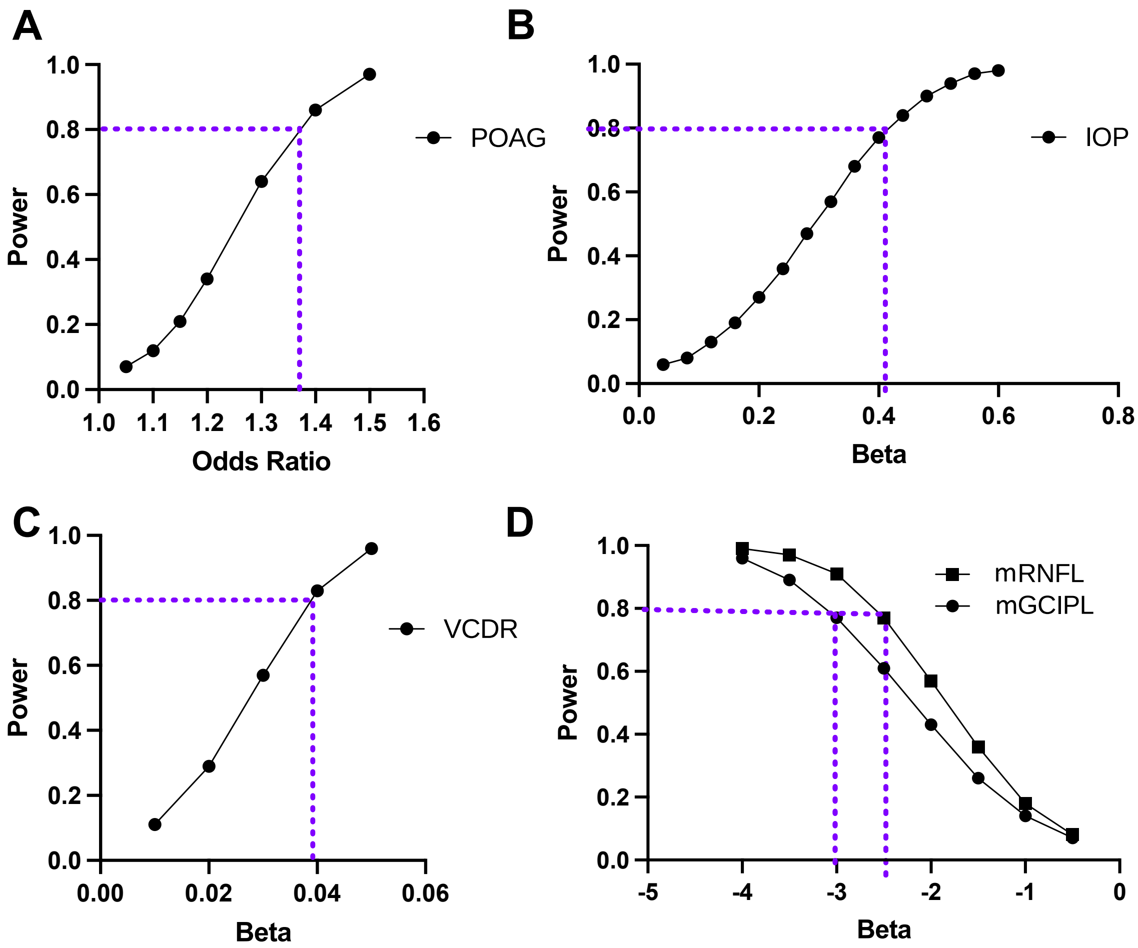


Supplementary Figure 2.

Graphs showing the power to detect a given odds ratio (panel A) and beta (panels B, C and D) for outcomes: POAG (n=216,257),[1] IOP (n=139,555),[2] VCDR (n=23,899),[3] RNFL and GCIPL (n=31,434).[4] Graphs are annotated to show the minimum odds ratio or beta for 80% power. We compared these to the published estimates of effect sizes where available, as well as our effect estimates. Data is presented for predicted direction of effect; increased 3-hydroxy-3-methylglutaryl-coenzyme A (HMG-CoA) reductase activity is hypothesized to be linked with increased POAG, IOP and VCDR and decreased mRNFL and mGCIPL thickness. These calculations showed that while we were well powered for detecting a significant effect in the VCDR outcome, we were less well powered for the other outcomes. See “Supplementary Methods for Mendelian Randomization Experiments” for references.

Abbreviations: IOP=intraocular pressure; mGCIPL= macular ganglion cell inner plexiform layer thickness in the macular region; mRNFL= retinal nerve fiber layer thickness in the macular region; POAG = primary open angle glaucoma; VCDR = vertical cup disc ratio.
