## Supplementary Table 1 for "Statin use in relation to intraocular pressure, glaucoma, and ocular coherence tomography parameters in the UK Biobank"

Characteristics of UK Biobank study population included in each analysis

| | Demographics |
|---|---|
| | **Mean (SD) or %** |
| ***Overall population (n=502,506; 2006-2010)*** | |
| Mean age, years (SD) | 56.5 (8.1) |
| Sex, % Women | 54.4 |
| Race/ethnicity, % Caucasian | 82.1 |
| ***IOP ANALYSES:*** | |
| ***IOP + statin + genetic data (n=118,153; 2009-2013)*** | |
| Mean age, years (SD) | 56.8 (8.0) |
| Sex, % Women | 53.5 |
| Race/ethnicity (% Caucasian) | 78.5 |
| Mean IOP, mmHg (SD) | 16.0 (3.8) |
| ***PREVALENT GLAUCOMA ANALYSES:*** | |
| ***Prevalent glaucoma + statin use + genetic data (n=192,283; 2006-2010)*** | |
| Mean age, years (SD) | 57.0 (8.0) |
| Sex (% Women) | 53.9 |
| Race/ethnicity (% Caucasian) | 91.7 |
| ***mRNFL ANALYSES:*** | |
| ***mRNFL + statin use + genetic data (n=41,638; 2009-2010)*** | |
| Mean age, years (SD) | 56.3 (8.1) |
| Sex, % Women | 52.7 |
| Race/ethnicity, % Caucasian | 90.8 |
| Mean mRNFL thickness, microns (SD) | 28.9 (3.8) |
| ***mGCIPL ANALYSES:*** | |
| ***mGCIPL + statin use + genetic data (n=41,547; 2009-2010)*** | |
| Mean age, years (SD) | 56.3 (8.1) |
| Sex, % Women | 52.8 |
| Race/ethnicity, % Caucasian | 90.8 |
| Mean GCIPL thickness, microns (SD) | 75.2 (5.2) |

Abbreviations: mGCIPL = ganglion cell inner plexiform layer; mRNFL = macula retinal nerve fiber layer thickness
