## Supplementary Table 2 for "Statin use in relation to intraocular pressure, glaucoma, and ocular coherence tomography parameters in the UK Biobank"

Characteristics of 118,153 UK Biobank participants with intraocular pressure measurements according to hypolipidemic use* at baseline (2006-2010)

| | **Non-users of hypolipidemics** (N = 94,255) | **Users of non-statin hypolipidemic medication only** (N = 3,305) | **Statin users** (N = 20,593) |
|---|---|---|---|
| **Age (year), mean (SD)** | 55.8 (8.1) | 59.4 (7.0) | 61.4 (6.1) |
| **Sex, n (%)** | 53,278 (56.5) | 2,069 (62.6) | 7,832 (38.0) |
| **Ethnicity, n (%)** | | | |
| *White British (Caucasian genetically)* | 73,618 (78.1) | 2,757 (83.4) | 16,318 (79.2) |
| *White (other)* | 11,482 (12.2) | 359 (10.9) | 2,121 (10.3) |
| *Black* | 3,033 (3.2) | 57 (1.7) | 493 (2.4) |
| *Asian (Indian/ Pakistani/ Bangladesh)* | 2,944 (3.1) | 58 (1.8) | 1,013 (4.9) |
| *Chinese* | 420 (0.4) | 11 (0.3) | 67 (0.3) |
| *Other* | 2,758 (2.9) | 63 (1.9) | 581 (2.8) |
| **Townsend Deprivation Index, mean (SD)** | -1.1 (3.0) | -1.5 (2.9) | -0.9 (3.1) |
| **Statin type, n (%)** | | | |
| *Simvastatin* | - | - | 14,602 (70.9) |
| *Atorvastatin* | - | - | 4,484 (21.8) |
| *Rosuvastatin* | - | - | 768 (3.7) |
| *Pravastatin* | - | - | 681 (3.3) |
| *Fluvastatin* | - | - | 54 (0.3) |
| *Multiple* | - | - | 4 (0.0) |
| **Non-statin hypolipidemic medication use, n (%)** | 0 (0.0) | 3,305 (100) | 1,474 (7.2) |
| **Serum total cholesterol (mmol/l), mean (SD)** | 5.9 (1.0) | 5.9 (1.0) | 4.7 (0.9) |
| **Serum triglyceride (mmol/l), mean (SD)** | 1.7 (0.9) | 1.7 (1.0) | 1.9 (1.0) |
| **Systolic blood pressure (mmHg), mean (SD)** | 136.4 (18.3) | 138.9 (19.1) | 141.4 (17.5) |
| **Systemic beta-blocker use, n (%)** | 2,765 (2.9) | 201 (6.1) | 4,255 (20.7) |
| **Diabetes, n (%)** | 2,084 (2.2) | 223 (6.7) | 4,763 (23.1) |
| **HbA1c (mmol/mol), mean (SD)** | 35.3 (5.1) | 36.3 (6.2) | 40.3 (9.7) |
| **Cardiovascular disease, n (%)** | 2,106 (2.2) | 187 (5.7) | 3,515 (17.1) |
| **Body mass index (kg/m$^2$), mean (SD)** | 26.9 (4.4) | 26.9 (4.4) | 29.0 (4.5) |
| **Physical activity (MET-hours/week), mean (SD)** | 44.7 (44.7) | 46.1 (42.3) | 40.4 (42.4) |
| **Smoking status, n (%)** | | | |
| *Never* | 54,576 (57.9) | 1,864 (56.4) | 9.712 (47.2) |
| *Past* | 30,742 (32.6) | 1,232 (37.3) | 8,731 (42.4) |
| *Current* | 8,937 (9.5) | 209 (6.3) | 2,150 (10.4) |
| **Alcohol drinking frequency, n (%)** | | | |
| *Never or special occasion only* | 18,794 (19.9) | 646 (19.5) | 4,945 (24.0) |
| *Ever and often* | 75,461 (80.1) | 2,659 (80.5) | 15,648 (76.0) |
| **Coffee (cups per day), mean (SD)** | 1.9 (1.7) | 1.8 (1.6) | 1.9 (1.8) |
| **Tea (cups per day), mean (SD)** | 3.1 (2.1) | 3.2 (2.0) | 3.1 (2.1) |
| **Spherical equivalent, mean (SD)** | -0.4 (2.7) | -0.2 (2.6) | 0.0 (2.6) |
| **IOP (mmHg), mean (SD)** | 15.9 (3.8) | 16.2 (3.9) | 16.3 (3.9) |
| **Prevalent glaucoma, n (%)** | 1,605 (1.7) | 74 (2.2) | 626 (3.1) |
| **Glaucoma MTAG GRS, mean (SD)** | 0 (1.0) | 0 (1.0) | 0 (1.0) |

Abbreviation: MTAG GRS = multi-trait analysis of genome wide association study genetic risk score

* “Statin users” were defined as anyone using statins (± other non-statin hypolipidemic drugs); while “users of non-statin hypolipidemic medication only” were defined as those not on any statins but using fibric acid agents, bile acid sequestrants, niacin-derivatives, cholesterol absorption inhibitors, omega-3 fats and other miscellaneous antihyperlipidemic agents
