## Supplementary Table 3 for "Statin use in relation to intraocular pressure, glaucoma, and ocular coherence tomography parameters in the UK Biobank"

Multivariable-adjusted association* between statin use and glaucoma related parameters in the UK Biobank among participants without prevalent glaucoma

| | **IOP (mmHg)** | **mRNFL (microns)** | **mGCIPL (microns)** |
|---|---|---|---|
| | (N = 116,287) | (N = 41,120) | (N = 41,029) |
| **Statin use** | Difference (95% CI); (p-value) n (sample size) | Difference (95% CI); (p-value) n (sample size) | Difference (95% CI); (p-value) n (sample size) |
| Non-user | Ref n=96,243 | Ref n=34,291 | Ref n=34,221 |
| User | 0.04 (-0.04, 0.11) (p=0.31) n=20,044 | -0.14 (-0.28, -0.01) (p=0.03) n=6,829 | -0.11 (-0.28, 0.07) (p=0.24) n=6,808 |

Abbreviations: mGCIPL=macular ganglion cell inner plexiform layer; mRNFL=macular retinal nerve fiber layer thickness; ref=reference

* Each model is adjusted for: age, $age^2$, sex, ethnicity (White, Black, Asian, and Other), deprivation, and spherical equivalent + non-statin hypolipidemic medication use, smoking status, number of cigarettes (only among current smokers), alcohol, physical activity + missing indicator, and coffee and tea intake, body mass index, systolic blood pressure, HbA1c + missing indicator, total serum cholesterol level + missing indicator and serum triglyceride level + missing indicator.
