## Supplementary Table 4 for "Statin use in relation to intraocular pressure, glaucoma, and ocular coherence tomography parameters in the UK Biobank"

Multivariable-adjusted association* between hypolipidemic drug use† and glaucoma related parameters in the UK Biobank

| | **IOP (mmHg)** | **Prevalent glaucoma** | **mRNFL (microns)** | **mGCIPL (microns)** |
|---|---|---|---|---|
| | (N = 118,153) | (N = 192,283) | (N = 41,638) | (N = 41,547) |
| **Hypolipidemic drug use** | Difference (95% CI); (p-value) n (sample size) | Odds Ratio (95% CI) (p-value) n (sample size) | Difference (95% CI) (p-value) n (sample size) | Difference (95% CI) (p-value) n (sample size) |
| Non-users of hypolipidemics | Ref n=94225 | Ref n=151118 | Ref n=33690 | Ref n=33,607 |
| Statin users | 0.05 (-0.02, 0.13) (p=0.16) n=20593 | 1.05 (0.98, 1.13) (p=0.15) n=34,951 | -0.13 (-0.27, 0.00) (p=0.05) n=6973 | -0.16 (-0.34, 0.01) (p=0.07) n=6955 |
| Users of non-statin hypolipidemics only | 0.005 (-0.14, 0.15) (p=0.95) n=3305 | 1.04 (0.92, 1.18) (p=0.55) n=6214 | 0.21 (-0.26, 0.67) (p=0.38) n=975 | -0.74 (-1.35, -0.13) (p=0.02) n=985 |

Abbreviations: mGCIPL=macular ganglion cell inner plexiform layer; mRNFL=macular retinal nerve fiber layer thickness; ref=reference

* Each model adjusted for age, age$^2$, sex, ethnicity (White, Black, Asian, and Other), deprivation, spherical equivalent, smoking status, number of cigarettes (only among current smokers), alcohol, physical activity + missing indicator, coffee and tea intake, body mass index, systolic blood pressure, diabetes, cardiovascular disease, HbA1C + missing indicator, baseline systemic beta-blocker use, serum total cholesterol + missing indicator, serum triglyceride + missing indicator

† "Statin users" were defined as anyone using statins (± other non-statin hypolipidemic drugs); while "users of non-statin hypolipidemic medication only" were defined as those not on any statins but using fibric acid agents, bile acid sequestrants, niacin-derivatives, cholesterol absorption inhibitors, omega-3 fats and other miscellaneous antihyperlipidemic agents
