## Supplementary Table 5 for "Statin use in relation to intraocular pressure, glaucoma, and ocular coherence tomography parameters in the UK Biobank"

Multivariable-adjusted analyses* assessing whether the glaucoma MTAG GRS modified the relation between statin use and the four glaucoma related outcomes or the relation between non-statin hypolipidemic drug use† and the four glaucoma outcomes in the UK BioBank

| | IOP (mmHg) | Prevalent glaucoma | mRNFL (microns) | mGCIPL (microns) |
|---|---|---|---|---|
| | (N = 118,153) | (N = 192,283) | (N = 41,638) | (N = 41,547) |
| | $Beta_{int}$ ($P_{int}$) | $OR_{int}$ ($P_{int}$) | $Beta_{int}$ ($P_{int}$) | $Beta_{int}$ ($P_{int}$) |
| **MTAG GRS*Statin use** | | | | |
| (GRS*Any statin use) | 0.05 (0.19) | 0.99 (0.81) | 0.05 (0.42) | -0.08 (0.31) |
| (GRS*Non-statin hypolipidemic medication use) | -0.04 (0.72) | 1.06 (0.64) | 0.15 (0.24) | 0.07 (0.70) |

Abbreviations: GRS = genetic risk score, int = interaction; MTAG = multi-trait association of genome wide association studies; OR = Odds Ratio;

* Model for IOP: adjusted for age, $age^2$, sex, ethnicity (White, Black, Asian, and Other), deprivation, spherical equivalent, smoking status, number of cigarettes (only among current smokers), alcohol, physical activity + missing indicator, coffee and tea intake, body mass index, systolic blood pressure, diabetes, cardiovascular disease (CVD), HbA1C + missing indicator, systemic beta-blocker use, serum total cholesterol (TC), serum triglycerides (TG) + missing indicator + TC*MTAG GRS + CVD*MTAG GRS + TG*MTAG GRS + age*MTAG GRS + $age^2$*MTAG GRS

Model for prevalent glaucoma: adjusted for age, $age^2$, sex, ethnicity (White, Black, Asian, and Other), deprivation, spherical equivalent + missing indicator, smoking status, number of cigarettes (only among current smokers), alcohol, physical activity + missing indicator, coffee and tea intake, BMI, SBP + missing indicator, diabetes, CVD, HbA1C, systemic beta-blocker use, TC, and TG + TC*MTAG GRS + CVD*MTAG GRS + TG*MTAG GRS + age*MTAG GRS + $age^2$*MTAG GRS

Model for mRNFL and mGCIPL: adjusted for age, $age^2$, sex, ethnicity (White, Black, Asian, and Other), deprivation, spherical equivalent, smoking status, number of cigarettes (only among current smokers), alcohol, physical activity + missing indicator, coffee and tea intake, BMI, SBP, diabetes, CVD, HbA1C + missing indicator, systemic beta-blocker use, TC and TG + missing indicator + TC*MTAG GRS + CVD*MTAG GRS + TG*MTAG GRS + age*MTAG GRS + $age^2$*MTAG GRS

† "Statin users" were defined as anyone using statins (± other non-statin hypolipidemic drugs); while "users of non-statin hypolipidemic medication only" were defined as those not on any statins but using fibric acid agents, bile acid sequestrants, niacin-derivatives, cholesterol absorption inhibitors, omega-3 fats and other miscellaneous antihyperlipidemic agents
