## Supplementary Table 6 for "Statin use in relation to intraocular pressure, glaucoma, and ocular coherence tomography parameters in the UK Biobank"

Mendelian Randomization analysis between 3-hydroxy-3-methylglutaryl-coenzyme A reductase activity and glaucoma-related outcomes*

| Outcome* | Method | Beta / Odds Ratio | 95% Confidence Interval | P-value |
|---|---|---|---|---|
| POAG | IVW | 0.86 | 0.52 - 1.41 | 0.54 |
| | Egger | 0.97 | 0.80 – 8.65 | 0.11 |
| | Egger intercept | NA | NA | 0.05 |
| | Weighted median | 0.79 | 0.60-1.05 | 0.11 |
| IOP (mmHg) | IVW | 0.07 | -0.26 – 0.41 | 0.67 |
| | Egger | -0.01 | -1.05 -1.03 | 0.98 |
| | Egger intercept | NA | NA | 0.86 |
| | Weighted median | 0.09 | -0.19 – 0.36 | 0.54 |
| VCDR | IVW | 0.04 | -0.02 – 0.09 | 0.19 |
| | Egger | 0.09 | -0.03 – 1.64 | 0.14 |
| | Egger intercept | NA | NA | 0.34 |
| | Weighted median | 0.04 | 0.01 – 0.06 | 0.02 |
| mRNFL (microns) | IVW | 0.70 | -0.24 -1.64 | 0.14 |
| | Egger | 0.004 | -4.81 - 4.82 | 1.00 |
| | Egger intercept | NA | NA | 0.76 |
| | Weighted median | 0.62 | -0.14 -1.38 | 0.11 |
| mGCIPL (microns) | IVW | 0.47 | -0.76 – 1.70 | 0.45 |
| | Egger | 0.68 | -3.08 – 4.43 | 0.72 |
| | Egger intercept | NA | NA | 0.91 |
| | Weighted median | 0.36 | -0.63 -1.35 | 0.47 |

Abbreviations: GWAS=genome wide association study; IGGC=International Glaucoma Genetics Consortium; IOP=intraocular pressure; IVW=inverse variance weighted; mRNFL=macular retinal nerve fiber layer; mGCIPL=macular ganglion cell inner plexiform layer; NA=not applicable; POAG=primary open angle glaucoma; VCDR=vertical cup disc ratio;.

*For the IOP dataset, we used 139,555 participants from three cohorts: UK Biobank, EPIC-Norfolk and the previously reported combined results from 14 European studies in the IGGC.

For the POAG dataset, we used the stage 1 GWAS summary data from a meta-analysis that combined 18 POAG studies in Europeans (16,677 POAG cases vs 199,580 controls).

The VCDR GWAS was performed as part of the metanalysis by the IGGC, combining multiple studies and included $n$ = 23,899 participants of European descent.

The mRNFL and mGCIPL GWAS used data solely from UK Biobank: OCT data that fit quality control measures was extracted from the densest populated well-mixed population within the overall dataset (n= 31434).
