## Supplementary Table 7 for "Statin use in relation to intraocular pressure, glaucoma, and ocular coherence tomography parameters in the UK Biobank"

Multivariable-adjusted association* between statin use and glaucoma related parameters in the UK Biobank among participants with total serum cholesterol <200mg/dl and no cardiovascular disease, diabetes or systemic beta blocker use

| | **IOP (mmHg)** | **Prevalent glaucoma** | **mRNFL (microns)** | **mGCIPL (microns)** |
|---|---|---|---|---|
| | (N = 26,639) | (N = 42,654) | (N = 9,434) | (N = 9,423) |
| **Statin use** | Difference (95% CI); (p-value) n (sample size) | Odds Ratio (95% CI); (p-value) n (sample size) | Difference (95% CI); (p-value) n (sample size) | Difference (95% CI); (p-value) n (sample size) |
| Non-user | Ref n=19,965 | Ref n=31,919 | Ref n=7,066 | Ref n=7,046 |
| User | 0.02 (-0.12, 0.15) (p=0.81) n=6,674 | 1.02 (0.90, 1.16) (p=0.75) n=10,735 | -0.16 (-0.39, 0.07) (p=0.17) n=2,368 | -0.42 (-0.72, -0.12) (p=0.01) n=2,372 |

Abbreviations: mGCIPL=macular ganglion cell inner plexiform layer; mRNFL=macular retinal nerve fiber layer thickness; ref=reference

* Model for IOP: adjusted for: age, $age^2$, sex, ethnicity (White, Black, Asian, and Other), deprivation, spherical equivalent, non-statin hypolipidemic medication use, smoking status, number of cigarettes (only among current smokers), alcohol, physical activity + missing indicator, and coffee and tea intake, body mass index, systolic blood pressure, HbA1c + missing indicator, total serum cholesterol level, and serum triglyceride level + missing indicator.

Model for prevalent glaucoma: adjusted for: age, $age^2$, sex, ethnicity (White, Black, Asian, and Other), deprivation, spherical equivalent + missing indicator, non-statin hypolipidemic medication use, smoking status, number of cigarettes (only among current smokers), alcohol, physical activity + missing indicator, and coffee and tea intake, body mass index, systolic blood pressure, HbA1c + missing indicator, total serum cholesterol level, and serum triglyceride level + missing indicator.

Model for mRNFL and mGCIPL: adjusted for: age, $age^2$, sex, ethnicity (White, Black, Asian, and Other), deprivation, spherical equivalent, non-statin hypolipidemic medication use, smoking status, number of cigarettes (only among current smokers), alcohol, physical activity + missing indicator, and coffee and tea intake, body mass index, systolic blood pressure, HbA1c + missing indicator, total serum cholesterol level, and serum triglyceride level + missing indicator.
