## Supplementary Table 8 for "Statin use in relation to intraocular pressure, glaucoma, and ocular coherence tomography parameters in the UK Biobank"

Main multivariable-adjusted* associations between statin use and glaucoma-related outcomes among UK Biobank participants (n=41,444) who have complete data on all outcomes

| | | **Glaucoma-related outcomes** (N =41,444) | | | |
|---|---|---|---|---|---|
| | | **IOP (mmHg)** | **Prevalent glaucoma† (N case= 567)** | **mRNFL† (microns)** | **mGCIPL† (microns)** |
| **Statin use** | **N (sample size)** | Difference (95% CI); (p-value) | Odds Ratio (95% CI) (p-value) | Difference (95% CI); (p-value) | Difference (95% CI); (p-value) |
| Non-user | 34,461 | **Ref** | **Ref** | **Ref** | **Ref** |
| User | 6,983 | -0.02 (-0.14, 0.10) (p=0.74) | 1.14 (0.87, 1.50) (p=0.34) | -0.13 (-0.26, 0.00) (p=0.05) | -0.13 (-0.31, 0.04) (p=0.13) |
| *Simvastatin* | 5,115 | -0.03 (-0.17, 0.10) (p=0.64) | 1.20 (0.89, 1.61) (p=0.23) | -0.13 (-0.27, 0.02) (p=0.08) | -0.16 (-0.35, 0.02) (p=0.09) |
| *Atorvastatin* | 1,373 | -0.06 (-0.29, 0.17) (p=0.62) | 0.81 (0.48, 1.37) (p=0.43) | -0.10 (-0.34, 0.14) (p=0.42) | -0.10 (-0.42, 0.21) (p=0.52) |
| *Rosuvastatin* | 254 | -0.11 (-0.60, 0.38) (p=0.66) | 0.97 (0.33, 2.82) (p=0.95) | -0.11 (-0.62, 0.40) (p=0.68) | 0.01 (-0.67, 0.69) (p=0.97) |
| *Pravastatin* | 222 | -0.28 (-0.80, 0.25) (p=0.30) | 1.42 (0.55, 3.68) (p=0.47) | -0.24 (-0.79, 0.31) (p=0.40) | -1.09 (-1.81, -0.36) (p=0.003) |
| *Fluvastatin* | 18 | -0.77 (-2.61, 1.07) (p=0.41) | 5.08 (0.57, 4.56) (p=0.15) | -0.37 (-2.31, 1.57) (p=0.71) | -2.31 (-4.87, 0.25) (p=0.07) |

Abbreviations: mGCIPL=macular ganglion cell inner plexiform layer; mRNFL=macular retinal nerve fiber layer thickness; ref=reference

* Adjusted for age, age$^2$, sex, ethnicity (White, Black, Asian, and Other), deprivation, spherical equivalent, smoking status, number of cigarettes (only among current smokers), alcohol, physical activity + missing indicator, coffee and tea intake, body mass index, systolic blood pressure, diabetes, cardiovascular disease, HbA1c + missing indicator, systemic beta-blocker use, serum total cholesterol + missing indicator and serum triglyceride + missing indicator

† Additionally adjusted for IOP
